# Historic 1994 influenza vaccine cohorts reveal breadth of antibody and B cell responses towards three decades of future influenza A and B viruses

**DOI:** 10.1101/2025.07.22.25332020

**Authors:** Thi H. O. Nguyen, Isabelle J. H. Foo, Ruth A. Purcell, Hyon-Xhi Tan, Georgia Deliyannis, Wuji Zhang, Louise Carolan, A. Jessica Hadiprodjo, Howard H. Huang, Lilith F. Allen, Ruth R. Hagen, L. Carissa Aurelia, Hayley A. McQuilten, Louise C. Rowntree, Lukasz Kedzierski, Samuel H. Wilks, Matthew R. McKay, Gregory A. Tannock, Stephen J. Kent, Karen Laurie, Annette Fox, Steven Rockman, Lorena E. Brown, Amy W. Chung, Adam K. Wheatley, Katherine Kedzierska

## Abstract

Influenza vaccination is the best way to combat annual influenza epidemics, yet the breadth of vaccine-induced humoral immunity towards decades of future differentially-evolving influenza A and B viruses is unclear. Using historic 1994 influenza vaccination cohorts of young and older adults, we defined antibody responses elicited by 1994 vaccination against future influenza strains spanning three decades of differentially-evolving influenza A (FLUAV) and B (FLUBV) viruses. Quality of antibody responses together with vaccine-induced and cross-reactive B-cell memory responses were investigated. Vaccination increased antibody titers against all 1994 vaccine components (H1N1 A/Texas/36/1991, H3N2 A/Beijing/32/1992, Yamagata B/Panama/45/1990) in young adults, but not B/Panama/45/90 in older adults. Antibodies towards future H1N1 strains were detected across younger and older adults. Older adults, additionally displayed boosted responses towards A/Michigan/45/2015, related to the 2009 pandemic strain known to induce cross-reactive antibodies with 1918-like H1N1 viruses. Antibody responses towards future rapidly-evolving H3N2 strains were minimal. Prominent boosting against earlier B/Yamagata/16/1988 and future Yamagata-lineage strains were found across younger and older adults. Individuals who responded strongly to B/Panama/45/1990 also responded to future FLUBV strains from Yamagata and Victorian lineages. Systems serology revealed qualitative differences in antigen-antibody signatures between younger and older adults at baseline before 1994 vaccination, with serological features towards vaccine antigens overlapping post-vaccination. Older adults, however, comprised divergent antibody signatures against future antigens featuring mature IgA1 responses, while younger adults featured more naive IgM responses. To define cross-reactive B-cell responses, fluorescently-labelled recombinant HA-probes were generated for vaccine and future influenza strains. 1994 vaccination induced cross-reactive memory B-cells towards vaccine and future H1 and FLUBV strains, but minimal responses for H3. Our study provides key insights into the breadth of vaccine-induced humoral immunity towards future influenza viruses over 30-years of FLUAV and FLUBV evolution, including newly-emerging pandemic strains, and the need to optimize future vaccine strategies, especially for the rapidly-evolving H3N2.

## INTRODUCTION

Over the past 30 years, seasonal inactivated influenza vaccines (IIV) continue to be the most effective strategy to prevent influenza virus infection. The trivalent IIV covers two influenza A virus (FLUAV) strains (H1N1 and H3N2) and one influenza B virus (FLUBV) strain (Victoria or Yamagata lineage). IIVs, traditionally being egg-based vaccines, have since progressed to include quadrivalent vaccines including both influenza B lineages, cell-based vaccines for people aged ≥ 6 months, high dose vaccines for ≥ 60 years, and adjuvanted vaccines for ≥ 65 years. People aged ≥ 65 years respond poorly to IIV (*1, 2*) and are more susceptible to influenza illness requiring hospitalization and death, accounting for 50–70% and 70–85% of cases, respectively (*3*). Adjuvanted vaccines are recommended for older adults in preference to the standard dose since they can boost more potent B cell responses in this older population (*4, 5*). Due to the rapid evolution of H3N2, the H3N2 vaccine strain is often mismatched with the circulating strains (*6*). Therefore, vaccine efficacy against H3N2 strains is lowest compared to the slower evolving H1N1 and FLUBV strains (*6, 7*), with older adults being most affected (*2, 8*).

Antibody responses are typically measured serologically using a surrogate haemagglutinin (HA) inhibition (HAI) assay or microneutralisation assay detecting HA-specific antibodies. B cell responses include generation of plasmablasts and HA-specific memory B cells, which positively correlate with increases in HAI antibody titres following IIV vaccination (*9*). Studies demonstrated that pre-existing immunity (i.e. prior exposures to influenza virus infections and/or vaccinations) and age-related effects can impact the magnitude and cross-reactive potential of the antibody and B cell response to seasonal influenza vaccination (*8, 10–14*). However, the breadth of vaccine-induced humoral immunity towards decades of future influenza A and B viruses in both adults and older adults is less defined.

Using historic 1994 influenza vaccination cohorts of younger and older adults, we defined pre-vaccination antibody reactivity, and antibody responses elicited by 1994 influenza vaccination against future influenza virus strains spanning three decades of differentially evolving influenza subtypes, H1N1, H3N2 and FLUBV. We also investigated the quality of antibody responses by defining antibody isotype, subclass and effector function, as well as vaccine-boosted and cross-reactive B cell memory responses using contemporary methods such as systems serology and fluorescently-labelled recombinant HA-probes.

We found that vaccination induced and/or boosted antibody titers against 1994 vaccine viral strains in younger and older adults. Antibody responses towards future H1N1 and FLUBV strains were observed in both age groups, but not towards future H3N2 strains. Qualitative differences in vaccine-specific antigen-antibody signatures were observed between younger and older adults prior to 1994 vaccination, but greatly overlapped post-vaccination, reflecting high quality vaccine-driven antibody responses in both age groups. Post-vaccination antibody responses against future antigens, however, showed distinct signatures in older adults which featured mature IgA1 responses, while younger adults featured more naive IgM responses. Finally, 1994 vaccination induced cross-reactive memory B cell responses towards the vaccine and future strains for H1 and FLUBV, but minimal responses for H3, supporting our antibody landscape data.

Thus, using a unique 1994 vaccinated cohort, into the breadth of vaccine-induced humoral immunity towards future influenza viruses over 30-years of FLUAV and FLUBV evolution, including newly-emerging pandemic strains, and the need to optimize future vaccine strategies, especially for the rapidly evolving H3N2.

## RESULTS

### 1994 IIV study cohort

Our 1994 IIV cohort included 89 younger adults and 68 older adults vaccinated in 1994 with the trivalent IIV Fluvax (Fig. 1A). The vaccine was derived from H1N1 A/Texas/36/1991, H3N2 A/Beijing/32/1992 and FLUBV Yamagata B/Panama/45/1990 reference strains. Blood samples were collected at baseline and ∼28 days (d) following vaccination for plasma and PBMCs, then cryopreserved for 30 years. Young adults had a median age of 19 years (range 18-53) and were 46% female, while older adults had a median age of 69 years (range 60-75) and were 49% female (Fig. 1, B and C). Young adults were predominantly born in the 1970s (85% of young cohort) when influenza B viruses diverged into Yamagata and Victoria lineages (Fig. 1D). Older adults were mainly born in the 1920s (88% of older cohort) and were exposed to H1N1 from the 1918-1919 H1N1 prototypical pandemic virus until the late 1960s when H2N2 emerged. Interestingly, one 75-year-old female was born during the 1918-1919 H1N1 pandemic, which spread globally and killed > 40 million people (*15*). The 1918 H1N1 pandemic virus was antigenically similar to the most recent 2009 H1N1 influenza pandemic virus that our cohort was not exposed to (*16*).

**Fig. 1.**
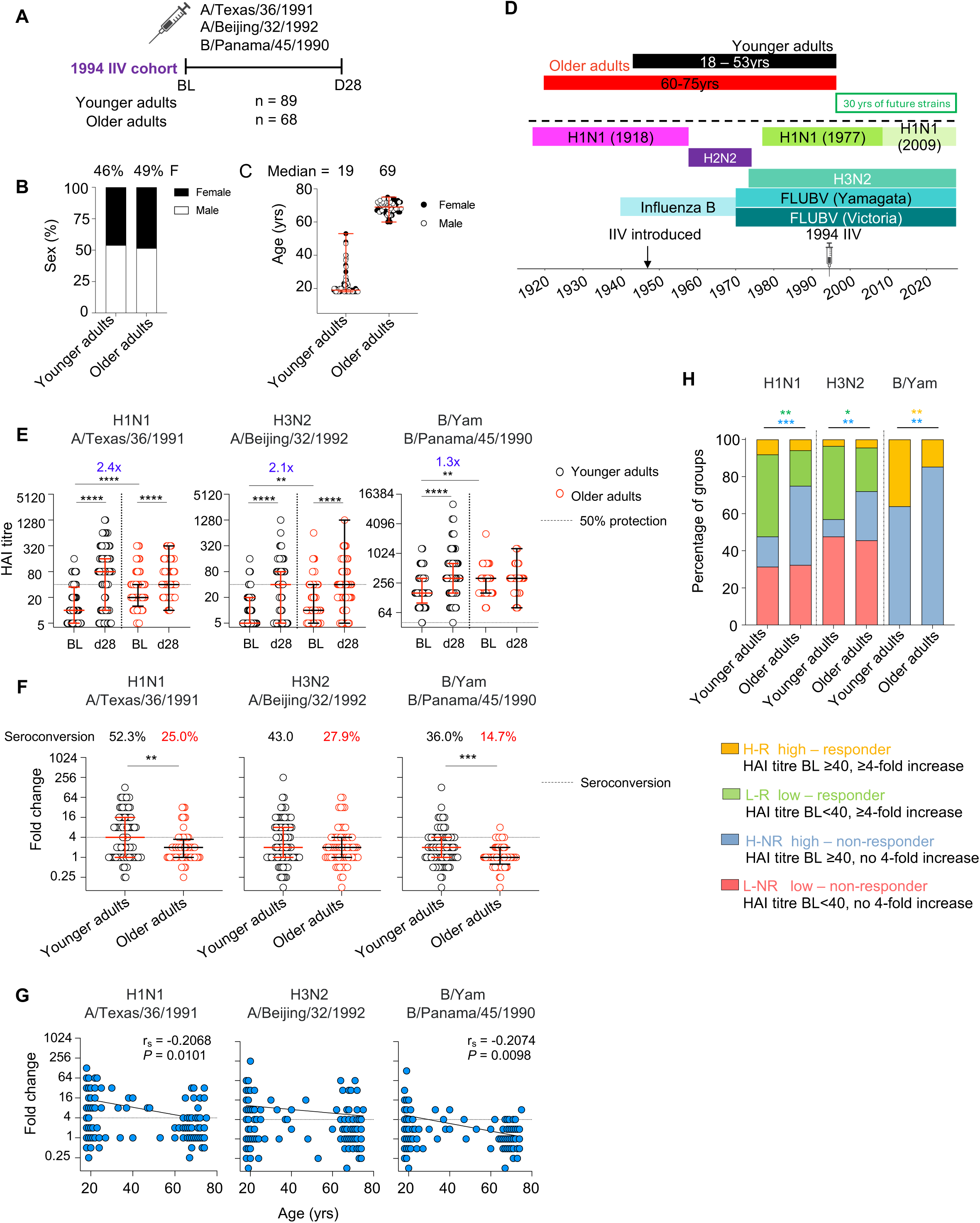
1994 IIV study cohort and vaccine responses. (**A**) 1994 IIV cohort sampling timepoints, (**B**) sex distribution and (**C**) age. (**D**) Exposure history of past and future circulating influenza virus strains. (**E**) HAI antibody titres towards 1994 IIV strains in young and older adults. (**F**) Fold-change HAI responses (D28/BL), where seroconversion is defined as 4 or more-fold change. (**E** and **F**) Median and IQR are shown. Statistical significance determined by Wilcoxon test for timepoint comparisons within an age group or Mann-Whitney for comparisons between adults and older adults. (**G**) Spearman’s correlation of fold-change HAI responses and age. (**H**) Frequency of four responder groups defined by baseline antibody titres and seroconversion status across strains and between age groups. Statistical significance was determined by two-sided chi-square test. \*\**P* < 0.01, \*\*\**P* < 0.001, \*\*\*\**P* < 0.0001.

### Antibody responses towards the 1994 IIV

To evaluate antibody responses towards the 1994 vaccine strains, haemagglutinin inhibition (HAI) assays were performed on the entire 1994 cohort. A HAI antibody titre of 40 has been found to represents 50% protection (*17*). Young adults had increased HAI antibody titres following vaccination to all three strains, while older adults only had increased HAI antibody responses to A/Texas/36/1991 and A/Beijing/32/1992. (Fig. 1E). However, older adults had 1.3-2.4-fold higher baseline HAI titres across all vaccine strains compared to younger adults. As a result, older adults had lower seroconversion rates (15-28%) compared to younger adults (36-52%) across all strains (Fig. 1F), where seroconversion describes a ≥ 4-fold increase in antibody titres from baseline levels. Magnitude of seroconversion negatively correlated with aging (Fig. 1G), which was more prominent for H1N1 (r_s_ =-0.2068) and B/Yam (r_s_ = - 0.2074) compared to H3N2 (r_s_ =-0.09021).

Cohorts were further divided into 4 groups based on their seroconversion status and whether their baseline antibody titres were below or above > 40 (Fig. 1H). “Low non-responders” were defined as individuals with baseline HAI titres < 40 and who did not seroconvert (< 4-fold increase). “High non-responders” had baseline HAI titres ≥ 40 and did not seroconvert. “Low responders” seroconverted with baseline HAI titres < 40 and “high responders” seroconverted with baseline HAI titres ≥ 40. We found that younger adults had higher proportions of low responders for H1N1 (44.2%, *P* = 0.001) and H3N2 (39.5%, *P* = 0.0352) compared to older adults (19.1% H1N1, 23.5% H3N2). Interestingly, older adults had higher proportions of high non-responders across all three strains (26.5-85.3%, *P* = 0.0003-0.0047) compared to younger adults (9.30-64.0%), suggesting that although they did not seroconvert, these older individuals still had immunity to the virus based on their seropositive status.

### Antibody responses towards future H1N1, H3N2 and FLUBV influenza virus strains

Previous studies have investigated how influenza vaccine antibody responses could back-boost previously circulating strains for H3N2(*13*). In our study, we assessed future-boosting in a subset of individuals (28 young and 28 older adults), measuring the 1994 vaccine-induced and/or boosted antibody responses towards future H1N1, H3N2, B/Yam and B/Vic influenza viruses included in the influenza vaccine over the past 3 decades (table S1), including the A/California/7/2009 H1N1 pandemic strain (Fig. 2A).

**Fig. 2.**
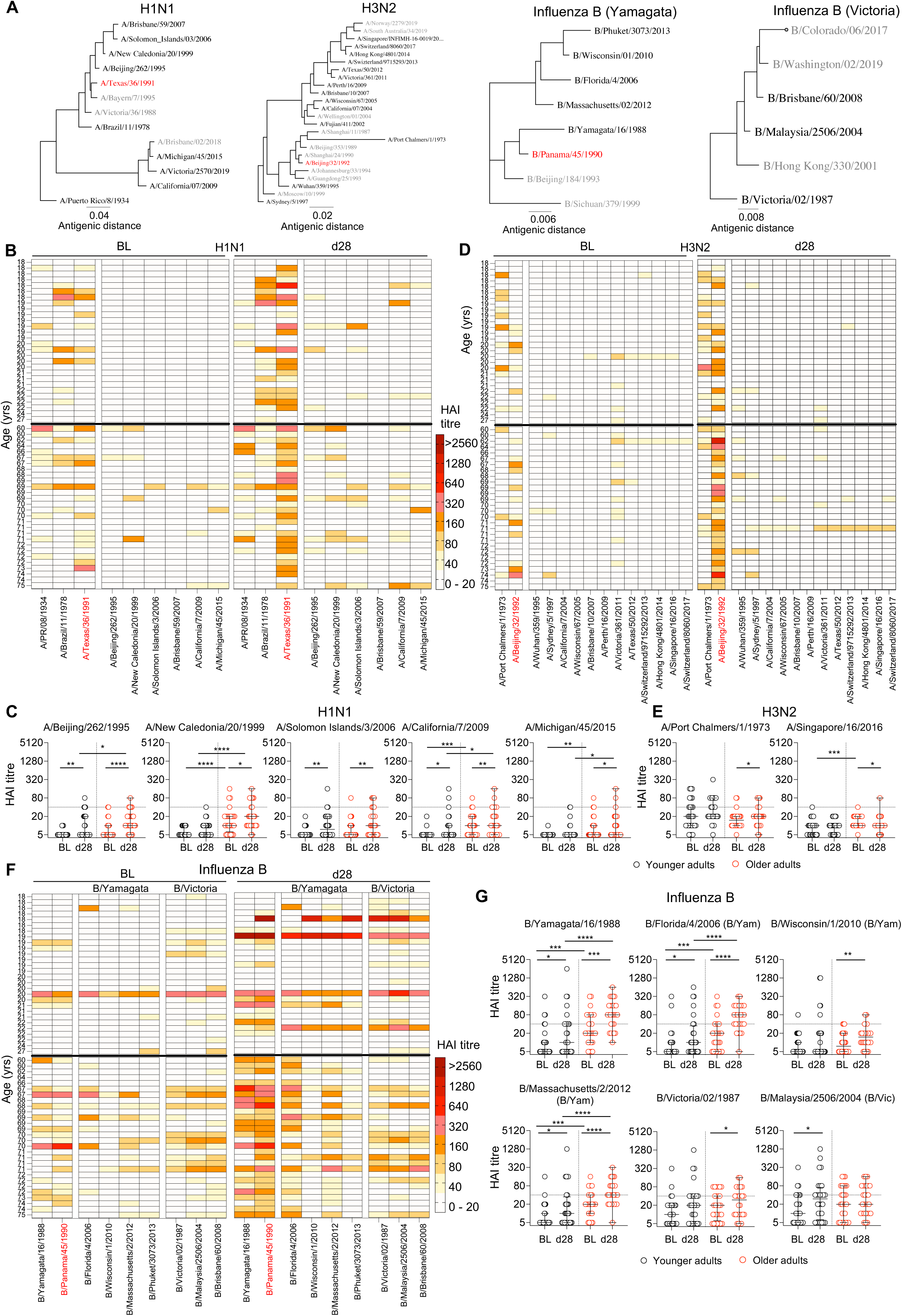
1994 IIV induces antibody titres against future H1N1 and FLUBV strains. (**A**) Phylogenetic tree of selected FLUAV and FLUBV reference strains. Red indicates 1994 IIV strains, black represent those selected for HAI testing on selected cohort of 28 young and 28 older adults. Heatmap of HAI antibody responses for each individual against (**B**) H1N1 and (**C**) H3N2 and (**F**) FLUBV strains. Representative HAI antibody graphs of (**D**) H1N1 and (**E**) H3N2 and (**G**) FLUBV strains. Median and IQR are shown. Statistical significance determined by Wilcoxon test for timepoint comparisons within an age group or by Mann-Whitney for comparisons between adults and older adults. \**P* < 0.05, \*\**P* < 0.01, \*\*\**P* < 0.001, \*\*\*\**P* < 0.0001.

We observed modest increases in HAI titres towards 3 future H1N1 strains (A/Beijing/262/1995, A/Solomon Islands/3/2006 and A/California/7/2009) in both younger and older adults following IIV (Fig. 2, B and C). However, older adults also had boosted antibody responses towards A/New Caledonia/20/1999 and A/Michigan/45/2015, the latter being closely related to the 2009 pandemic strain (Fig. 2A). Compared to younger adults, older adults had higher baseline antibody titres towards A/New Caledonia/20/1999, pandemic A/California/7/2009 and pandemic-like A/Michigan/45/2015 strains which increased following IIV (Fig. 2C), suggesting more cross-reactive antibody responses to future strains including pandemic-like strains in older adults. Three older individuals (aged 69, 71 and 75) were seropositive for A/California/7/2009 and one 70-year-old to A/Michigan/45/2015 at baseline (HAI = 40-80) (Fig. 2B), potentially having been exposed to the earlier 1918 pandemic strain, but only the eldest participant seroconverted against the pandemic strain following IIV. Although none of the younger adults were seropositive for A/California/7/2009 or A/Michigan/45/2015 at baseline (HAI < 40), three individuals did respond to either strain following IIV.

In contrast, antibody responses towards future H3N2 strains were minimal (Fig. 2D), reflecting rapid evolution of H3N2. Only vaccine-directed responses against the earlier A/Port Chalmers/1/1973 strain were observed in older adults (Fig. 2E). Prominent boosting against the earlier B/Yamagata/16/1988 and future B/Florida/4/2006 and B/Massachusetts/2/2012 Yamagata-lineage strains were observed in both adults and older adults following vaccination (Fig. 2, F and G), with older adults having higher baseline and d28 antibody levels compared to younger adults. Older adults also showed responses to future Yamagata-lineage B/Wisconsin/1/2010 as well as to previously circulating B/Victoria/02/1987 which was not evident for younger adults. Instead, boosting in younger adults was towards the future Victoria-lineage B/Malaysia/2506/2004 (Fig. 2G). Interestingly, individuals who responded strongly to the B/Panama/45/1990 vaccine strain, especially younger adults, could respond to multiple future FLUBV strains from both lineages.

Geometric mean titre (GMT) antibody landscapes within young and older adults strikingly showed how little boosting occurred against future H3N2 strains in comparison to robust future boosting against multiple future H1N1 and FLUBV strains following vaccination (Fig. 3), further highlighting the need to improve vaccine strategies for H3N2, especially for the elderly at risk of life-threatening influenza.

**Fig. 3.**
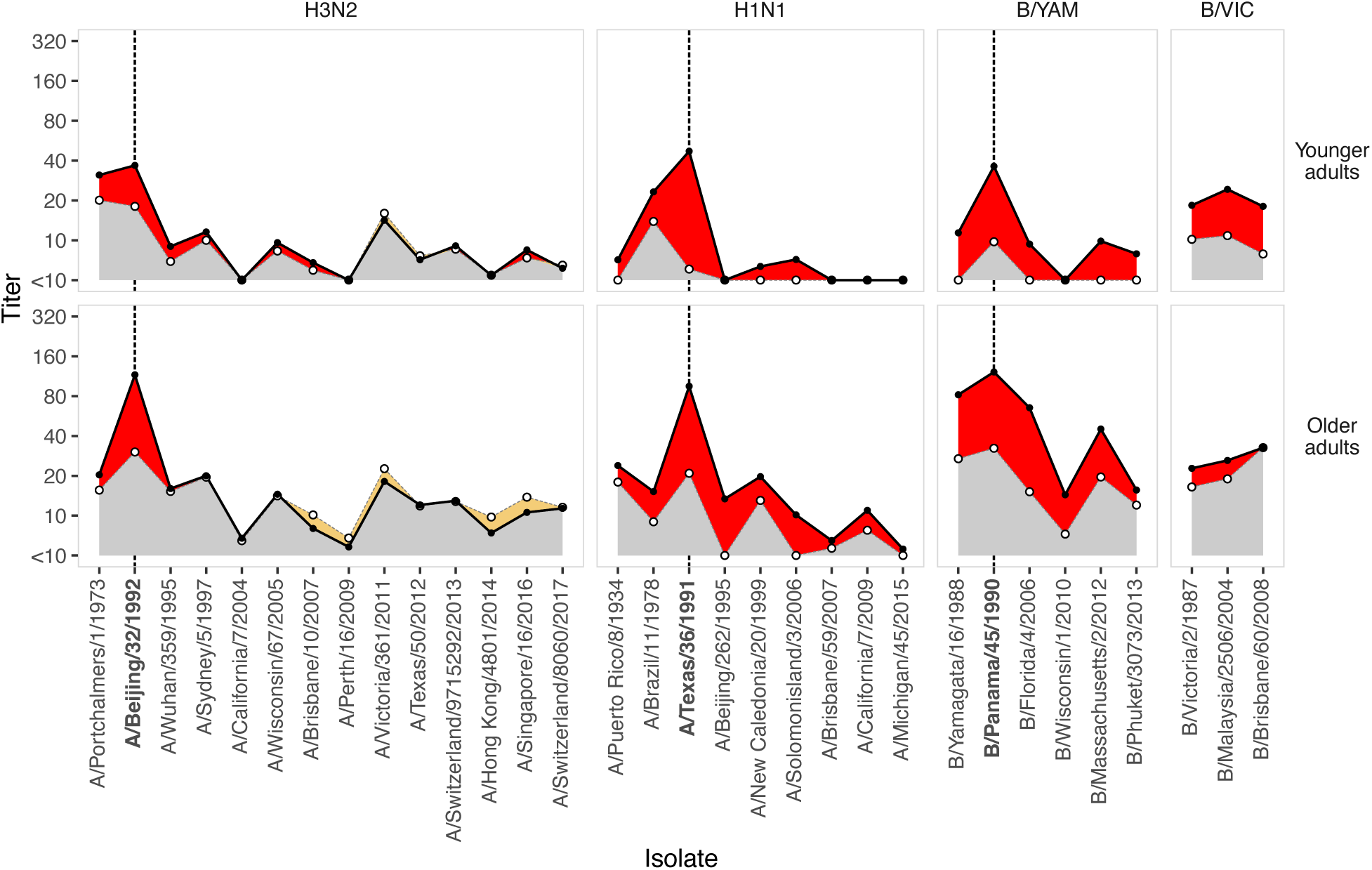
GMT landscapes of vaccine responses in the younger and older adult groups. Panels are split by age group and virus subtype. In each panel, the dashed grey line and open points show pre-vaccination GMTs for each isolate tested, while the solid black line and solid points show post-vaccination GMTs. Areas shaded red show an increase in GMT from pre-post-vaccination, beige shows a decrease, and grey shows the level of background reactivity found in both pre and post-vaccination samples. Dashed vertical black lines mark the vaccine strain for each subtype. Viruses are ordered by date of isolation on x-axis, giving an approximate genetic / antigenic progression.

### Divergent antibody response signatures between younger and older adults

The multidimensional systems serology approach can comprehensively define antibody responses including their isotype, subclass and effector function to a range of different antigens. We developed a multiplex antigen array that included 14 influenza antigens (8 HA, 3 neuraminidase (NA) and 3 nucleoprotein (NP) proteins) from the 1994 vaccine strains and future virus strains (fig. S1 and table S2) to characterise vaccine-induced and cross-reactive antibody responses. These experiments assessed plasma from 35 younger and 38 older adults, overlapping with those from the antibody landscape analyses. Each of the 14 influenza antigens were assessed for the presence of 12 different antibody responses including total IgG, IgM isotypes, Ig subclasses (IgG1, IgG2, IgG3, IgG4, IgA1) and Fc gamma receptors (FcγIIa-H131, FcγRIIa-R131, FcγRIIb, FcγRIIIa-V158 and FcγIIIa-F158, table S3), making up a total of 168 features. Antibody binding to Fc receptors on immune cells can lead to antibody-dependent cellular phagocytosis (FcγRIIa, i.e. CD32a) and antibody-dependent cell-mediated cytotoxicity (FcγIIIa, i.e. CD16) of target cells.

Changes in antibody expression levels to each influenza antigen were measured, comparing baseline and d28 post-vaccination within each age group (fig. S2 to S5). Differences in MFI responses were summarised by volcano plots to compare between age groups as well as between timepoints, which included i) all antigens, ii) 1994 vaccine antigens (H1-HA A/Texas/36/1991 and H3-HA A/Beijing/32/1992), or iii) post-1994 antigens in the analyses (Fig. 4, A and B, fig. S6 and table S2). When comparing differences between baseline versus d28 post-vaccination, most responses increased post-vaccination for both age groups, with only a few IgM responses enriched at baseline (fig. S6, A to C). Analyses of features between age groups at either timepoints revealed dominant skewing of responses towards older adults, particularly at baseline and against future antigens, whereas minimal enrichment of features was observed in older adults when only the 1994 vaccine antigens were analysed (Fig. 4, A and B, and fig. S6, D and E).

**Fig. 4.**
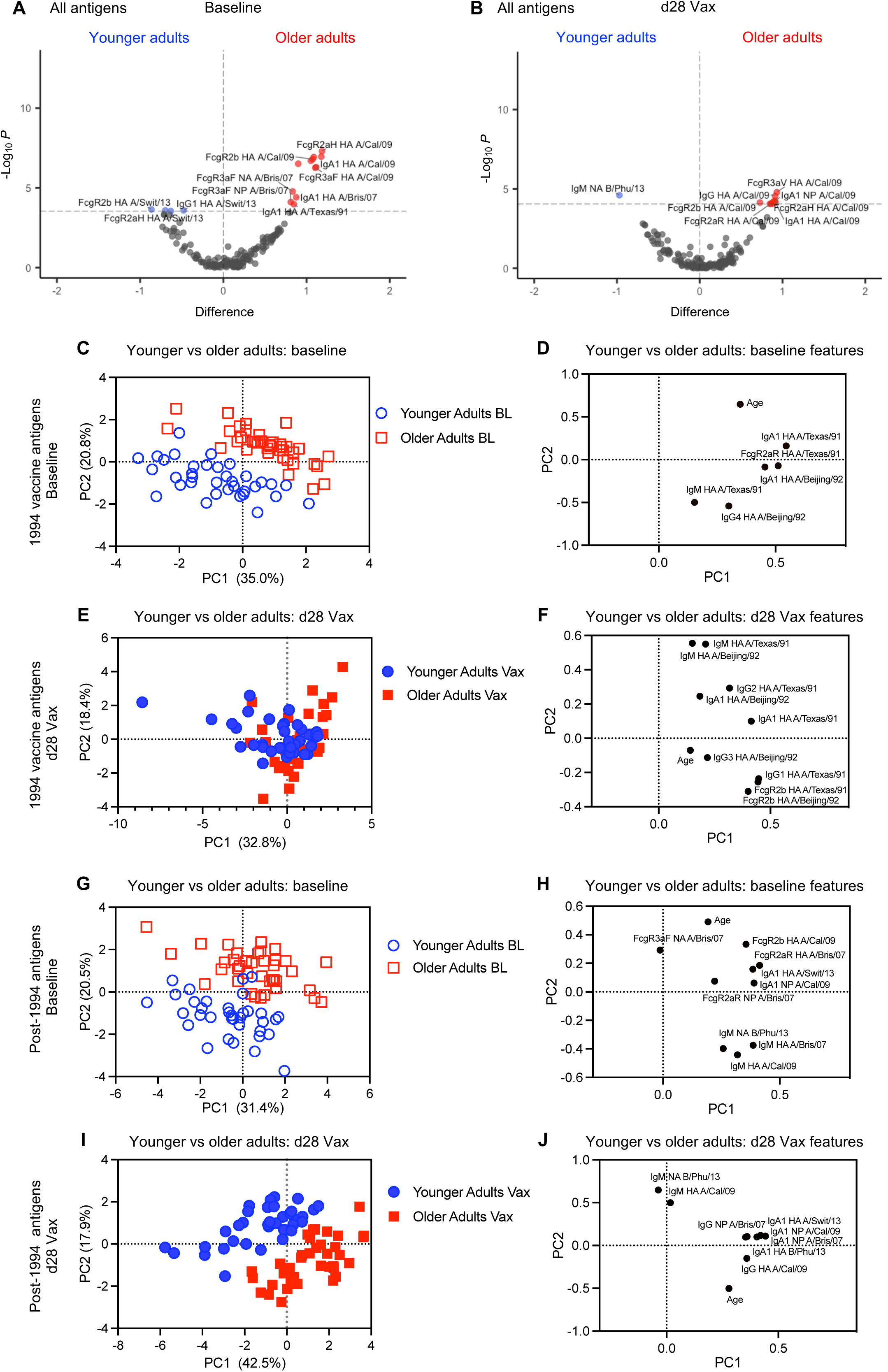
Multidimensional systems serology of antibody responses to vaccine and future viral strains. (**A** and **B**) Volcano plots of antigen-antibody features between young and older adults at (**A**) baseline and (**B**) d28 post-vaccination timepoints. Z-score differences in Log_10_(MFI+1) for each variable from baseline to d28 post-vaccination were determined by Wilcoxon matched-pairs signed rank test, and between age cohorts by Wilcoxon rank-sum/Mann-Whitney U test. The Holm correction for multiple comparisons was made; unadjusted-Log_10_ *P* values are plotted, with the horizontal threshold line plotted at an adjusted (-Log_10_) *P* value of 0.05. PCA of feature selected data from dataset including 1994 vaccine antigens plus age as variable features comparing (**C**) scores and (**D**) features of antibody responses for young and older adults at baseline and (**E**) scores and **(F**) features at d28 post-vaccination. 1994 vaccine antigens included H1-HA A/Texas/36/1991 and H3-HA A/Beijing/32/1992. (**G**) PCA scores and (**H**) antibody features from post 1994 vaccine antigens plus age as comparing young and older adults at baseline and (**I** and **J**) post-vaccination.

To further probe the qualitative differences between age groups, LASSO (least absolute shrinkage and selection operator) regression and Principal Component Analysis (PCA) were performed on the above dataset including age as a feature variable. For the 1994 vaccine antigens analysis, pre-vaccination baseline antibody responses revealed two distinct clusters between younger and older adults (Fig. 4C), which separated diagonally across PC2 based on selection of 6 out of 73 features (Fig. 4D). Older adults had stronger baseline Fc-receptor and IgA1 responses to the vaccine antigens, indicating a more mature baseline immune profile prior to vaccination, whereas younger adults exhibited IgM responses characteristic of naïve antibody responses. Post-vaccination responses against vaccine antigens were reduced to 10 features and generated only partial clustering between younger and older adults across PC1 (Fig. 4, E and F), analogous to the volcano plot analyses above, suggesting that immunity to the vaccine antigens were quite similar between age groups.

Baseline responses against future antigens were selected from 10 out of 145 variable features revealing clear clustering of age groups diagonally across PC2 (Fig. 4, G and H), which was very similar to their baseline responses against vaccine antigens. Strikingly, post-vaccination responses against future antigens revealed clear clustering between young and older adults diagonally across PC2 (Fig. 4, I and J), which was distinct from the 1994 vaccine antigens post-vaccination responses where partial clustering was observed (Fig. 4, E and F). This future antigen analysis suggested a more divergent cross-reactive signature in older adults which featured mature IgA1 responses, while younger adults featured more naive IgM responses (Fig. 4, I and J), reflective of their baseline signatures prior to vaccination (Fig. 4, G and H). These divergent signatures between younger and older adults were also reflected in our analyses including all 168 variable features plus age at both timepoints (fig. S7, A to D). Analyses between timepoints showed partial clustering across PC1 for both age groups with almost all features associated with post-vaccination responses (fig. S7, E to H), supporting our volcano plot analyses.

Taken together, our systems serology data revealed qualitative differences occurring prior to vaccination between younger and older adults in response to vaccine antigens and future antigens. Vaccine-specific responses greatly overlapped post-vaccination, representing high quality vaccine-induced antibody responses in both age groups. In contrast, post-vaccination responses against future antigens were quite distinct, with older adults exhibiting more mature immune responses, perhaps reflective of immune imprinting or biases due to longer history of prior exposures.

### 1994 IIV-driven cross-reactive memory B cells

To define influenza-specific B cell memory responses, fluorescently-labelled recombinant HA-probes were custom generated for H1, H3 and influenza B HA from the 1994 vaccine strains (fig. S8). To detect cross-reactive responses against future strains, HA-probes representing two future vaccine strains for each subtype were included (fig. S9 and table S3). These were H1-HA-probes from A/Brisbane/59/07 (2009 IIV) and A/Victoria/2570/2019 (H1N1)pdm09-like (2021-2022 IIV), H3-HA-probes from A/Perth/16/2009 (2010-2012 IIV) and A/Switzerland/9715292/2013 (2015 IIV), and FLUBV-HA-probes from B/Brisbane/60/2008 (2010-2012, 2016-2017 IIV) and B/Phuket/3073/2013 (2015, 2018-2025 IIV).

After 30 years of cryopreservation, viability of these historic PBMC samples were extremely high averaging 89-92% for both groups (Fig. 5A). For the 1994 vaccine strains, H1-HA Texas^+^ memory B cells increased 5.2-fold in young adults following vaccination (*P* < 0.0001), which trended 1.4-fold higher in older adults albeit not significantly (Fig. 5BC). Vaccine-driven Texas^+^ frequencies were also higher in younger adults compared to older adults (*P* = 0.0458). H3-HA Beijing^+^ memory B cells responses did not generally increase for both groups. However, FLUBV-HA Panama^+^ memory B cells responses robustly increased in both younger and older adults following vaccination (3.8-fold and 2.3-fold, respectively) with older adults having higher baseline frequencies compared to younger adults (*P* < 0.0001).

**Fig. 5.**
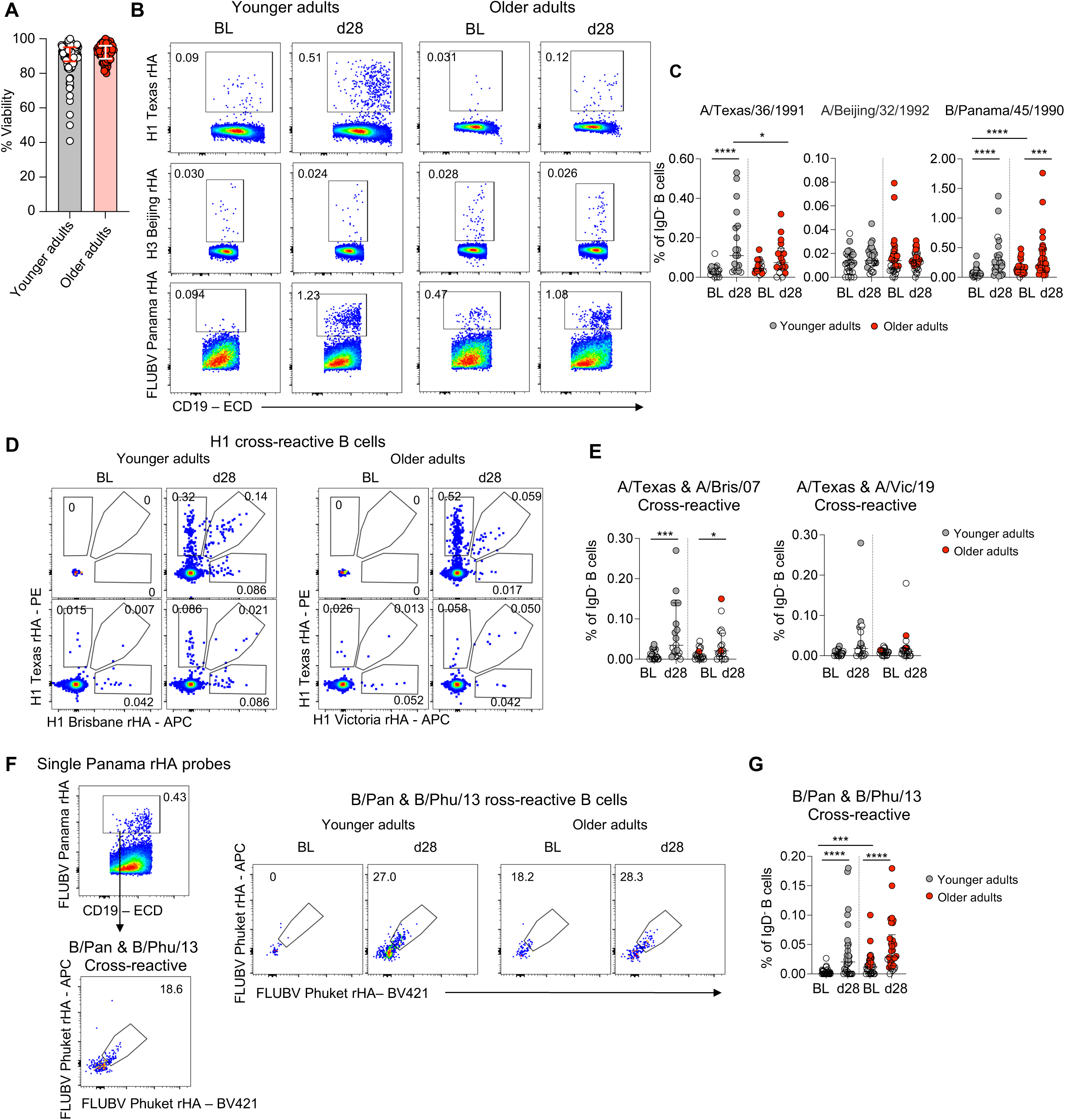
**1994 IIV-boosted cross-reactive influenza-specific B cell memory responses**. (**A**) Viability of 30-year cryopreserved PBMC samples. (**B**) Representative HA-probe staining for 1994 IIV strain-specific memory B cells. (**C**) Frequency of 1994 IIV strain-specific IgD^-^ B cells at baseline and d28 post-vaccination in adults and older adults. (**D**) Representative staining and (**E**) frequency of cross-reactive H1-specific B cell responses. (**F**) Representative staining and (**G**) frequency of cross-reactive B/Yamagata-specific B cell responses. Medians and IQRs are shown. Open circles depict <5 events. Statistical significance determined by Wilcoxon test for timepoint comparisons within an age group or by Mann-Whitney for comparisons between adults and older adults. \**P* < 0.05, \*\*\**P* < 0.001, \*\*\*\**P* < 0.0001.

To detect cross-reactive H1-specific memory B cell responses, H1-HA probes were initially split into two panels of Texas^+^Bris/07^+^ and Texas^+^Vic/19^+^ (Fig. 5D). Further optimisation of H3-HA and FLUBV-HA probes allowed us to include 3 probes in a single staining panel (fig. S10 and table S3). Although at lower frequencies, we observed increased responses in H1-cross-reactive Texas^+^Bris/07^+^ memory B cells in both younger and older adults following vaccination (*P* = 0.0010 and *P* = 0.0290, respectively), and increased trends for the Texas^+^Vic/19^+^ memory B cell response albeit not significant (Fig. 5E). Patterns were reflected in total Bris/07^+^ and Vic/19^+^ memory B cell responses (fig. S10, A and B). With less H3-HA Beijing^+^ memory B cell responses (10-fold lower than H1-Texas^+^ and FLUBV-Panama^+^ responses), we did not detect any cross-reactive H3-specific B cells. Rather, single-positive H3-Perth^+^ memory B cells were detected in both groups at baseline but did not in general increase following vaccination (fig. S10, C and D). We did observe, however increased responses in H3-Switz^+^ memory B cells in younger adults (*P* = 0.0011, 3.1-fold), whereas older adults already had higher baseline levels of H3-Switz^+^ memory B cells, without any further significant increases following vaccination (1.8-fold) (fig. S10, C and D).

Cross-reactive FLUBV-HA-specific memory B cells responses towards the Yamagata lineage strains were highly observed, with increases in Panama^+^Phuket^+^ memory B cells in both young (10.9-fold) and older adults (2.9-fold) following vaccination (Fig. 5, F and G). This was reflected in the total Phuket^+^ memory B cell response (fig. S10, E and F). FLUBV Bris^+^ memory B cells from the Victoria lineage were detected in both age groups but did not increase following vaccination (fig. S10, E and F) or cross-react with the Yamagata probes.

Phenotype (CD21 and CD27 expression) and isotype composition of total B cells did not change following vaccination, with the exception of young adults who had a small decrease in CD21^-^CD27^-^ B cells (Fig. 6A). At baseline and d28, younger adults had more IgD^+^ B cells and less IgM^+^ B cells compared to older adults (Fig. 6B). Whereas, for HA-specific B cells, we observed prototypical increases in the proportion of the activated memory CD21^-^CD27^+^ population for H1-Texas^+^ and FLUBV-Panama^+^ following vaccination, which coincided with a reduction in the resting memory CD21^+^CD27^+^ population (Fig. 6C). Cross-reactive H1 and FLUBV responses also showed similar activation patterns following vaccination, significant for the Panama^+^Phuket^+^ memory B cell response from older adults (Fig. 6D) and single probe analyses for H1-Bris/07^+^, H1-Vic/19^+^ and FLUBV-Phuket^+^ memory B cells (fig. S10G). Although not cross-reactive to the H3-Beijing vaccine probe, boosted H3-Switz^+^ memory B cells also showed increased activation phenotype in both age groups (fig. 10G). In terms of isotype, HA-specific B cells were predominantly IgG^+^ and did not change following vaccination (Fig. 6, E and F, and fig. S10H).

**Fig. 6.**
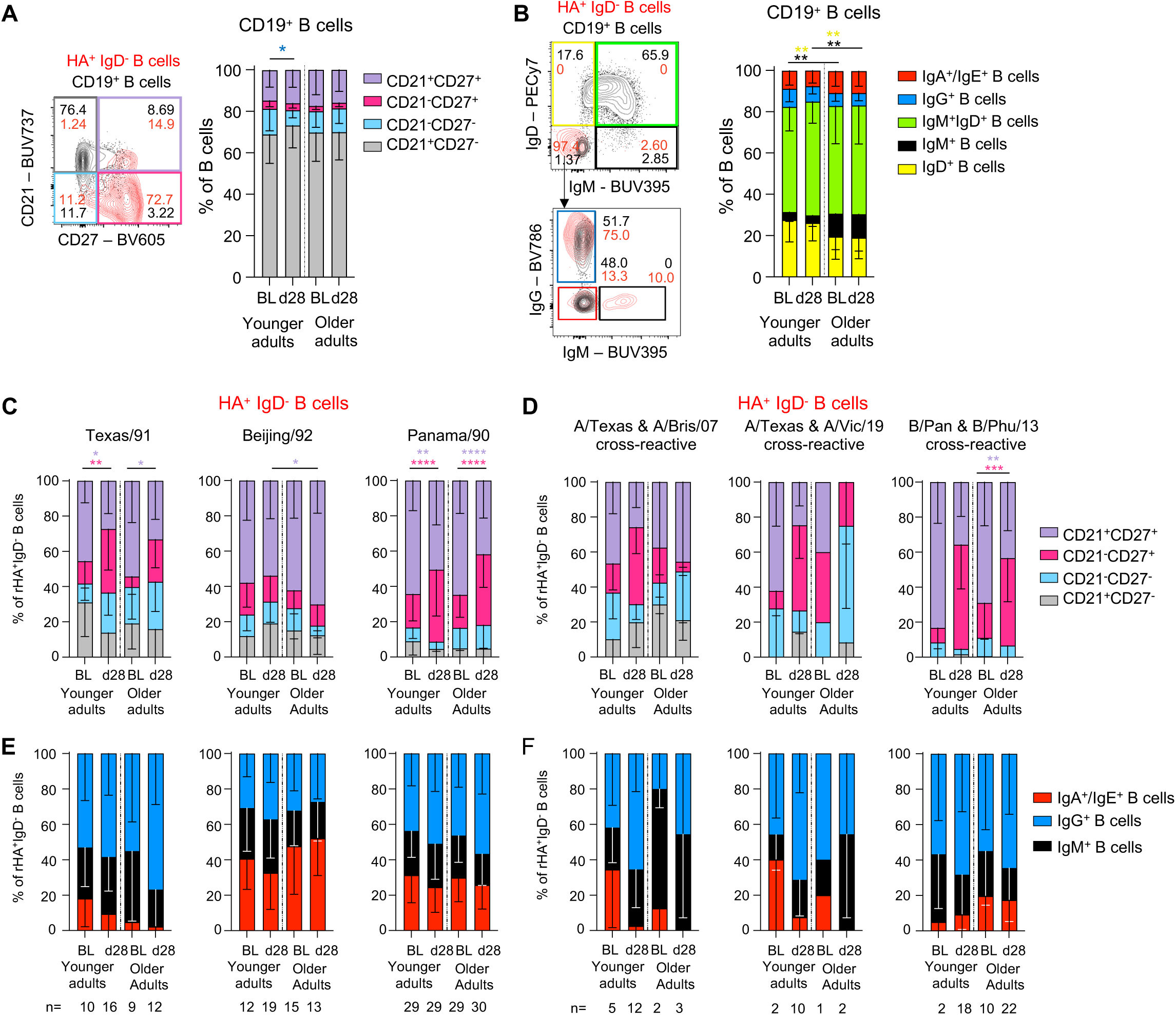
Phenotype of 1994 IIV-strain specific and cross-reactive memory B cell responses. (**A**) Phenotypic and (**B**) isotypic gating of total CD19^+^ B cells and HA^+^ IgD^-^ B cells. Graphs of total B cells are shown. (**C**) Phenotype and (**E**) isotype of 1994 IIV strain-specific memory B cells. (**D**) Phenotype and (**F**) isotype of cross-reactive H1 and FLUBV memory B cells. Mean and SD are shown. Statistical significance determined by Tukey’s multiple comparisons test. \**P* < 0.05, \*\**P* < 0.01, \*\*\**P* < 0.001, \*\*\*\**P* < 0.0001.

Our findings thus provide evidence for boosting and generation of vaccine-specific and cross-reactive memory B cell responses, particularly for H1 and FLUBV future strains. The change in activation phenotype, but not isotype following vaccination, represents a prototypical response in agreement to our previous study comparing influenza-specific memory B cell responses following influenza virus infection versus vaccination (*18*).

## DISCUSSION

In our study, we aimed to define pre-existing antibody responses elicited by influenza vaccination against future influenza virus strains. We analysed unique and historic influenza vaccination cohorts of young and older adults from 1994 and assessed future immune responses spanning three decades of differentially evolving influenza subtypes, H1N1, H3N2 and FLUBV. Our main findings showed that following vaccination, antibody responses increased against all three 1994 vaccine strains in young adults, but only H1N1 and H3N2 in older adults; that high-quality vaccine-induced antibody responses were achieved in both age groups, but divergent signatures existed against future antigens; and that antibody responses and cross-reactive memory B cell responses increased against future H1N1 and FLUBV strains but were less evident for H3 strains.

In our 1994 cohort study of young and older adults, we used birth year to estimate their exposure history and examined whether vaccination could elicit antibody responses against future H1N1, H3N2 and FLUBV strains spanning 3 decades including the 2009 pandemic and pandemic-like H1N1 strains (A/California/7/2009 and A/Michigan/45/2015). Our antibody landscape data revealed modest but significantly increased responses to multiple H1 and FLUBV future strains in both young and older adults following 1994 vaccination. However, antibody titres from older adults did fall above the seropositive threshold level more often than those from younger adults at both baseline and post-vaccination. Supporting the antibody landscape data, both age groups showed robust increases in vaccine-probe^+^ cross-reactive memory B cell responses against H1N1 A/Brisbane/59/2007 and B/Phuket/3073/2013, and in some individuals, increased cross-reactive responses against the A/Victoria/2570/2019 (H1N1)pdm09-like strain were observed. Although antibody responses against A/Brisbane/59/2007 were below the seropositivity threshold for both age groups, presence of H1N1-Texas^+^Brisbane^+^ memory B cells revealed that vaccination can also generate cross-reactive memory B cell responses targeting non-neutralising epitopes.

Our multiplex data provide a very sensitive approach to measure the quality of antibody responses against the vaccine and future antigens. One limitation was that there were no FLUBV/Panama antigens commercially available for the 1994 antigen analyses. However, our data suggested clustering was more impacted by Ig subtype and Fc effector function, and less so by the antigen sub-strain. For example, older adults had features of IgA1 and FcγRII antibody responses representing more mature responses while younger adults had features of naïve IgM responses. This was observed against all antigens prior to vaccination and was a continuing feature against future antigens post-vaccination suggesting a more cross-reactive signature in older adults. Conversely, post-vaccination responses against the vaccine antigens greatly overlapped representing high quality vaccine-specific responses in both age groups.

We had four older participants who were born in the late 1910s and early 1920s that may have been infected as a child with the 1918 H1N1 pandemic strain, which closely resembles the 2009 pandemic strain. Interestingly, these were the only participants seropositive for A/California/7/2009 and/or A/Michigan/45/2015 at baseline (HAI of 40 and above) with the eldest (75 years old) having a boosted A/California/7/2009 response, whereas none of the younger adults were seropositive for these future strains at baseline. Vaccination did however generate antibody responses against A/California/7/2009 and/or A/Michigan/45/2015 in three younger adults above the positivity threshold, which coincided with high antibody responses towards the vaccine A/Texas/36/1991 strain. This suggests that in some individuals, vaccination can elicit cross-reactive antibody responses between the vaccine subtype strain and future strains. These results corroborate previous vaccination studies demonstrating that influenza vaccination can induce broadly cross-reactive neutralising antibodies against both current and historical subtypes (*19, 20*). Similarly, mRNA COVID-19 boosters can elicit strong cross-reactive antibody and memory B cell responses against spike proteins from antigenically distinct SARS-CoV-2 variants that emerged post-vaccine distribution (*21, 22*).

We provide evidence of cross-reactive antibody responses between the two FLUBV lineages in both young and older adults who were potentially exposed to either lineage before the 1980s. In our 1994 vaccine study, our younger adults, born mainly in the early 1970s, had less exposure to FLUBV than our older adults, born in the 1920s, who experienced FLUBV strains since the 1940s. Both young and older adults robustly responded to the vaccine B/Panama/45/1990 Yamagata-lineage strain. However, older adults had much higher baseline and d28 antibody levels to the ancestral B/Yamagata/16/1988 and future Yamagata strains (B/Florida/4/2006 and B/Massachusetts/2/2012) compared to younger adults who were less seropositive for these strains. Young adults who responded strongly to the B/Panama/45/1990 vaccine component were able to respond to multiple future FLUBV strains from both lineages. Recent investigation by Edler *et al*. (*14*) revealed that birth cohorts between 1980s to late 1990s, when Yamagata strains predominated, exhibited highest antibody titres against future Yamagata-lineage strains, which were 2-fold higher than antibody levels against future Victoria-lineage strains. Whereas earlier birth cohorts (1940–1980) demonstrated cross-reactivity to future Victoria-lineage strains (*14*).

In our 1994 cohorts, we could not detect any cross-reactive Yamagata-Victoria lineage Panama^+^Brisbane^+^ or Phuket^+^Brisbane^+^ memory B cell responses, only cross-reactive Yamagata-lineage Panama^+^Phuket^+^ memory B cell responses or single-positive Victoria-lineage Brisbane^+^ memory B cells, including in the young participants who had antibody responses to both lineages. Whereas, we have previously shown that cohorts sampled during the mid-2010s have highly cross-reactive antibody and memory B cell responses to both lineages (*23*).

Our colleagues have previously defined the impact of prior influenza virus infection in shaping the H3N2 vaccination response in a 2016 IIV cohort study (*10*). They found that in healthy adults, people with at least one H3N2 infection had on average 2-3-fold higher antibody titres against the vaccine strain compared to those that had no prior H3N2 infection (*10*). Vaccinees with prior infection had higher and preserved antibody titres against the future H3N2 strain that was circulating a year later. Importantly, only 1.4% of those with prior infection became ill due to the H3N2 virus circulating one year after vaccination, compared to 14% with no prior infection, suggesting that vaccine efficacy was enhanced by pre-existing immunity by way of previous infection exposures. Given the impact of pre-existing immunity, they concluded that vaccination induced more of a recalled antibody-mediated response, rather than the production of de novo antibody responses from the naïve B cell pool (*10*).

Although our vaccine-specific H3-Beijing-probe showed no cross-reactive staining with the 2 future H3 probes, we could detect single-positive H3-Perth^+^ and H3-Switzerland^+^ memory B cells either at baseline or post-vaccination with their respective probes. We did observe an increase in the H3-Switzerland^+^ memory B cell response following vaccination suggesting activation of a B cell memory response, particularly in the younger adults. We could, however, detect single-positive H3-Beijing^+^ memory B cells, albeit at 10-fold lower frequencies, which did not increase following vaccination.

Thus, using our unique 1994 vaccinated cohort, we provide evidence for boosting of vaccine-specific and cross-reactive memory B cell responses, particularly for H1 and FLUBV future strains. Our study provides key insights into understanding how influenza vaccines protect against future influenza viruses, including newly emerging pandemic strains, and the need to optimize future vaccine strategies, especially for H3N2.

## MATERIALS AND METHODS

### Study design

The 1994 cohort was established as part of a Phase IV study to evaluate immune responses to the IIV (FLUVAX^®^, CSL Limited, Melbourne Australia) in a younger adult cohort (*n* = 89, 18-53 years) and an older adult cohort (*n* = 68, 60-75 years). Blood was collected at day 0 prior to vaccination and between days 24-32 post-vaccination. Serum was stored at-20°C or below for serological assays. PBMCs were cryopreserved at-196°C for cellular assays. All participants provided written informed consent. Study was approved by the Royal Melbourne Institute of Technology Ethics Committee (#40/93 and #41/93) and the University of Melbourne Human Research Ethics Committee (#13344 and #31236).

### Viruses used in HAI assays

Viruses for HAI assays were obtained from WHO Collaborating Centre for Reference and Research on Influenza (Melbourne, Australia) and CSL Seqirus Ltd (Parkville, Australia). List of viruses are detailed in table S1.

### Antibody landscapes

HAI antibody landscapes against the vaccine strain and other influenza strains from table S1 were measured at the WHO Collaborating Centre for Reference and Research on Influenza. When calculating geometric mean titers (GMTs) for pre and post-vaccination titers in younger and older adult groups, a maximum-likelihood approach to account for nondetectable titers (e.g. <10) was used as described (*24*), and implemented using the “gmt” function from the titertools R package (*25*).

### Multiplex bead array assay

Plasma samples were diluted 1:200 in PBS before incubating 1:1 (final dilution 1:400) with influenza proteins (HA, NA, NP) coupled to magnetic carboxylated beads (table S2), as described (*26*). The optimal plasma dilution was validated by correlation of EC_50_ responses with responses at the chosen single dilution and selected for suitability across detectors and antigens (fig. S1). SIV gp120-and Tetanus-coupled beads were included as negative and positive controls, respectively. The following day, samples were incubated with soluble FcγR dimers or mouse anti-human detector antibodies that were either PE-conjugated or biotinylated, followed by incubation with streptavidin-PE for biotinylated detection reagents (table S4), before reading on an Intelliflex Luminex instrument system (Luminex, Austin, TX, USA), as described (*26*). Samples were run in duplicate. For multivariate analysis, data (y) was log-transformed (y = log10(x+1)) before being normalised by mean centring and variance scaling of each feature. LASSO (least absolute shrinkage and selection operator) regression and Principal Component Analysis (PCA) were performed and visualized using MATLAB (MathWorks, Natick, USA), as described(*27, 28*). Frequency of selected samples was considered as the criteria of variable importance. Resampling of data was performed via 10-fold cross-validation and repeated 1,000 times (*26*). Figures were graphed using prism. Volcano plots were plotted in R version 4.4.1 using the EnhancedVolcano package (v1.22.0). Median fluorescence intensities (MFI) were right shifted by 1, then Log_10_ transformed using the formula Log_10_(MFI + 1), and z score scaled for each analysis comparison. Differences (z score group A - z score group B) between each feature from pre-to post-vaccination were determined by Wilcoxon matched-pairs signed rank test, and between age cohorts by Wilcoxon rank-sum/Mann-Whitney U test, using the rstatix package (v0.7.2). The Holm correction for multiple comparisons was made; unadjusted-Log_10_ p values are plotted, with the horizontal threshold line plotted at an adjusted (-Log_10_) p value of 0.05.

### HA-specific B cell responses

Vaccine-boosted and cross-reactive memory B cell responses were measured by flow cytometry (fig. S8 and S9) using a panel of in-house fluorescently labelled recombinant HA-probes comprising the 1994 IIV strain and two future strains for H1, H3 and FLUBV groups. PBMC samples were split evenly across the three groups. PBMCs were thawed, stained with HA probes and antibody panels as described in table S3 and table S5, then fixed with 1% PFA and acquired immediately on a LSR Fortessa (BD Biosciences), as described (*9, 18*). For H1N1, samples were split into two staining panels of A/Texas/36/1991 and A/Brisbane/59/2007 recombinant HA-probes, and A/Texas/36/1991 and A/Victoria/2570/2019 (H1N1)pdm09-like recombinant HA-probes. For H3 and FLUBV panels, all three probes were included in one staining panel.

### Statistical analysis

Statistical significance of nonparametric datasets (two-tailed) was determined using GraphPad Prism v10 software. Mann-Whitney U-test (unpaired) and Wilcoxon sign-rank test (paired) was used for comparisons between two groups. Tukey’s multiple comparison test was performed to compare row means between more than two groups.

### List of Supplementary Materials

**Fig. S1.** Validation of multiplex bead array assay.

**Fig. S2.** H1N1 responses across IgG subtypes.

**Fig. S3.** H1N1 responses across Fcγ receptors.

**Fig. S4.** H3N2 responses across IgG subtypes and Fcγ receptors.

**Fig. S5.** FLUBV responses IgG subtypes and Fcγ receptors.

**Fig. S6.** Volcano plots of antigen-antibody features from systems serology analyses.

**Fig. S7.** PCA analyses of young and older adults.

**Fig. S8.** FACS gating strategies to measure probe-specific memory B cell responses.

**Fig. S9.** Validation of recombinant H1, H3, and IBV HA probes on donors who received the IIV in 2022.

**Fig. S10.** Phenotype and isotype of future virus strain-specific memory B cell responses.

**Table S1.** List of influenza virus strains used in the study.

**Table S2.** Influenza antigens used for the multiplex assay.

**Table S3.** Fluorescently labelled recombinant HA-specific probes.

**Table S4.** Mouse anti-human antibody detectors used in multiplex assay.

**Table S5.** Mouse anti-human antibody panel for measuring HA-specific B cells.

## Data Availability

All study data are available upon reasonable request to the authors.

## Data Availability

All data produced in the present study are available upon reasonable request to the authors and are available in the main text or the supplementary materials.

## Acknowledgments

The 1994 cohort study was originally sponsored by CSL Limited. We thank all the participants who were involved in the study.

## Funding

This research was funded in whole or part by the National Health and Medical Research Council Investigator Grants: EL1 to THON (#1194036), EL1 to H-XT (#2009711), EL1 to LCR (#2026357), L3 to SJK (#2016491), EL2 to AWC (#2008092), L1 to AKW (#2026762) and L2 to KK (#2033783). MRM was supported by an Australian Research Council Future Fellowship (FT200100928) and Discovery Project (DP230102850). For the purposes of open access, the author has applied a CC BY public copyright licence to any Author Accepted Manuscript version arising from this submission. We thank the Melbourne Cytometry Platform for their services.

## Author Contributions

KK led the study. KK, THON, AWC and AKW supervised the study. THON, IJHF, RAP, H-XT, WZ, HHH, LCR, LK, KL, AF, AWC, AKW and KK designed the experiments. THON, IJHF, RAP, H-XT, WZ, LC, AJH, HHH and LFA performed and analysed experiments. RRH, LCA, HAM, SHW, MRM and AWC analysed data. H-XT, SJK, KL, SR, AF, AWC and AKW provided crucial reagents. GD, GAT, SR and LEB established the 1994 cohort. THON, KK, IJHF, RAP and AWC wrote the manuscript. All authors reviewed and approved the manuscript.

## Competing interests

All authors declare no competing interests.

## Data and materials availability

All data are available in the main text or the supplementary materials.

## Notes

### Competing Interest Statement

The authors have declared no competing interest.

### Author Declarations

All participants provided written informed consent. Study was approved by the Royal Melbourne Institute of Technology Ethics Committee (#40/93 and #41/93) and the University of Melbourne Human Research Ethics Committee (#13344 and #31236).

## REFERENCES AND NOTES

1. A. C. Henry, N. Y. Zheng, M. Huang, A. Cabanov, K. T. Rojas, K. Kaur, S. F. Andrews, A. E. Palm, Y. Q. Chen, Y. Li, K. Hoskova, H. A. Utset, M. C. Vieira, J. Wrammert, R. Ahmed, J. Holden-Wiltse, D. J. Topham, J. J. Treanor, H. C. Ertl, K. E. Schmader, S. Cobey, F. Krammer, S. E. Hensley, H. Greenberg, X. S. He, P. C. Wilson, Influenza Virus Vaccination Elicits Poorly Adapted B Cell Responses in Elderly Individuals. Cell Host Microbe 25, 357–366 e356 (2019).

2. S. Zhu, J. Quint, T. Leon, M. Sun, N. J. Li, C. Yen, M. W. Tenforde, B. Flannery, S. Jain, R. Schechter, C. Hoover, E. L. Murray, Estimating Influenza Vaccine Effectiveness Against Laboratory-Confirmed Influenza Using Linked Public Health Information Systems, California, 2023-2024 Season. J Infect Dis, (2025).

3. Centers for Disease Control and Prevention (CDC). vol. 2025.

4. C. A. DiazGranados, A. J. Dunning, M. Kimmel, D. Kirby, J. Treanor, A. Collins, R. Pollak, J. Christoff, J. Earl, V. Landolfi, E. Martin, S. Gurunathan, R. Nathan, D. P. Greenberg, N. G. Tornieporth, M. D. Decker, H. K. Talbot, Efficacy of high-dose versus standard-dose influenza vaccine in older adults. N Engl J Med 371, 635–645 (2014).

5. A. Domnich, L. Arata, D. Amicizia, J. Puig-Barbera, R. Gasparini, D. Panatto, Effectiveness of MF59-adjuvanted seasonal influenza vaccine in the elderly: A systematic review and meta-analysis. Vaccine 35, 513–520 (2017).

6. J. R. Chung, M. A. Rolfes, B. Flannery, P. Prasad, A. O’Halloran, S. Garg, A. M. Fry, J. A. Singleton, M. Patel, C. Reed, t. I. H. S. N. Us Influenza Vaccine Effectiveness Network, I. S. D. C. f. D. C. the Assessment Branch, Prevention, Effects of Influenza Vaccination in the United States During the 2018-2019 Influenza Season. Clin Infect Dis 71, e368–e376 (2020).

7. S. G. Sullivan, M. B. Chilver, K. S. Carville, Y. M. Deng, K. A. Grant, G. Higgins, N. Komadina, V. K. Leung, C. A. Minney-Smith, D. Teng, T. Tran, N. Stocks, J. E. Fielding, Low interim influenza vaccine effectiveness, Australia, 1 May to 24 September 2017. Euro Surveill 22, (2017).

8. F. Liu, F. L. Gross, S. N. Jefferson, C. Holiday, Y. Bai, L. Wang, B. Zhou, M. Z. Levine, Age-specific effects of vaccine egg adaptation and immune priming on A(H3N2) antibody responses following influenza vaccination. J Clin Invest 131, (2021).

9. M. Koutsakos, A. K. Wheatley, L. Loh, E. B. Clemens, S. Sant, S. Nüssing, A. Fox, A. W. Chung, K. L. Laurie, A. C. Hurt, S. Rockman, M. Lappas, T. Loudovaris, S. I. Mannering, G. P. Westall, M. Elliot, S. G. Tangye, L. M. Wakim, S. J. Kent, T. H. O. Nguyen, K. Kedzierska, Circulating TFH cells, serological memory, and tissue compartmentalization shape human influenza-specific B cell immunity. Science Translational Medicine 10, eaan8405 (2018).

10. M. Auladell, H. V. M. Phuong, L. T. Q. Mai, Y. Y. Tseng, L. Carolan, S. Wilks, P. Q. Thai, D. Price, N. T. Duong, N. L. K. Hang, L. T. Thanh, N. T. H. Thuong, T. T. K. Huong, N. T. N. Diep, V. T. N. Bich, A. Khvorov, L. Hensen, T. N. Duong, K. Kedzierska, D. D. Anh, H. Wertheim, S. D. Boyd, K. L. Good-Jacobson, D. Smith, I. Barr, S. Sullivan, H. R. van Doorn, A. Fox, Influenza virus infection history shapes antibody responses to influenza vaccination. Nat Med 28, 363–372 (2022).

11. S. F. Andrews, Y. Huang, K. Kaur, L. I. Popova, I. Y. Ho, N. T. Pauli, C. J. Henry Dunand, W. M. Taylor, S. Lim, M. Huang, X. Qu, J. H. Lee, M. Salgado-Ferrer, F. Krammer, P. Palese, J. Wrammert, R. Ahmed, P. C. Wilson, Immune history profoundly affects broadly protective B cell responses to influenza. Sci Transl Med 7, 316ra192 (2015).

12. S. F. Andrews, K. Kaur, N. T. Pauli, M. Huang, Y. Huang, P. C. Wilson, High preexisting serological antibody levels correlate with diversification of the influenza vaccine response. J Virol 89, 3308–3317 (2015).

13. J. M. Fonville, S. H. Wilks, S. L. James, A. Fox, M. Ventresca, M. Aban, L. Xue, T. C. Jones, N. M. H. Le, Q. T. Pham, N. D. Tran, Y. Wong, A. Mosterin, L. C. Katzelnick, D. Labonte, T. T. Le, G. van der Net, E. Skepner, C. A. Russell, T. D. Kaplan, G. F. Rimmelzwaan, N. Masurel, J. C. de Jong, A. Palache, W. E. P. Beyer, Q. M. Le, T. H. Nguyen, H. F. L. Wertheim, A. C. Hurt, A. Osterhaus, I. G. Barr, R. A. M. Fouchier, P. W. Horby, D. J. Smith, Antibody landscapes after influenza virus infection or vaccination. Science 346, 996–1000 (2014).

14. P. Edler, L. S. U. Schwab, M. Aban, M. Wille, N. Spirason, Y. M. Deng, M. A. Carlock, T. M. Ross, J. A. Juno, S. Rockman, A. K. Wheatley, S. J. Kent, I. G. Barr, D. J. Price, M. Koutsakos, Immune imprinting in early life shapes cross-reactivity to influenza B virus haemagglutinin. Nat Microbiol 9, 2073–2083 (2024).

15. K. R. Short, K. Kedzierska, C. E. van de Sandt, Back to the Future: Lessons Learned From the 1918 Influenza Pandemic. Front Cell Infect Microbiol 8, 343 (2018).

16. R. A. Medina, B. Manicassamy, S. Stertz, C. W. Seibert, R. Hai, R. B. Belshe, S. E. Frey, C. F. Basler, P. Palese, A. Garcia-Sastre, Pandemic 2009 H1N1 vaccine protects against 1918 Spanish influenza virus. Nat Commun 1, 28 (2010).

17. T. K. Tsang, S. Cauchemez, R. A. Perera, G. Freeman, V. J. Fang, D. K. Ip, G. M. Leung, J. S. Malik Peiris, B. J. Cowling, Association between antibody titers and protection against influenza virus infection within households. J Infect Dis 210, 684–692 (2014).

18. T. H. O. Nguyen, M. Koutsakos, C. E. van de Sandt, J. C. Crawford, L. Loh, S. Sant, L. Grzelak, E. K. Allen, T. Brahm, E. B. Clemens, M. Auladell, L. Hensen, Z. Wang, S. Nussing, X. Jia, P. Gunther, A. K. Wheatley, S. J. Kent, M. Aban, Y. M. Deng, K. L. Laurie, A. C. Hurt, S. Gras, J. Rossjohn, J. Crowe, J. Xu, D. Jackson, L. E. Brown, N. La Gruta, W. Chen, P. C. Doherty, S. J. Turner, T. C. Kotsimbos, P. G. Thomas, A. C. Cheng, K. Kedzierska, Immune cellular networks underlying recovery from influenza virus infection in acute hospitalized patients. Nat Commun 12, 2691 (2021).

19. M. G. Joyce, A. Wheatley, P. Thomas, G.-Y. Chuang, C. Soto, R. Bailer, A. Druz, I. Georgiev, R. Gillespie, M. Kanekiyo, W.-P. Kong, K. Leung, S. Narpala, M. Prabhakaran, E. Yang, B. Zhang, Y. Zhang, M. Asokan, J. Boyington, T. Bylund, S. Darko, C. Lees, A. Ransier, C.-H. Shen, L. Wang, J. Whittle, X. Wu, H. Yassine, C. Santos, Y. Matsuoka, Y. Tsybovsky, U. Baxa, J. Mullikin, K. Subbarao, D. Douek, B. Graham, R. Koup, J. Ledgerwood, M. Roederer, L. Shapiro, P. Kwong, J. Mascola, A. McDermott, Vaccine-Induced Antibodies that Neutralize Group 1 and Group 2 Influenza A Viruses. Cell 166, 609–623 (2016).

20. F. Krammer, The human antibody response to influenza A virus infection and vaccination. Nature Reviews Immunology 19, 383–397 (2019).

21. M.-C. Trieu, A. Reynaldi, W. S. Lee, H.-X. Tan, A. Kelly, R. Esterbauer, R. J. Cox, J. Audsley, J. Sasadeusz, D. S. Khoury, M. P. Davenport, D. Cromer, A. K. Wheatley, S. J. Kent, J. A. Juno, Bivalent mRNA booster vaccination recalls cellular and antibody immunity against antigenically divergent SARS-CoV-2 spike antigens. npj Vaccines 10, 74 (2025).

22. R. R. Goel, S. A. Apostolidis, M. M. Painter, D. Mathew, A. Pattekar, O. Kuthuru, S. Gouma, P. Hicks, W. Meng, A. M. Rosenfeld, S. Dysinger, K. A. Lundgreen, L. Kuri-Cervantes, S. Adamski, A. Hicks, S. Korte, D. A. Oldridge, A. E. Baxter, J. R. Giles, M. E. Weirick, C. M. McAllister, J. Dougherty, S. Long, K. D’Andrea, J. T. Hamilton, M. R. Betts, E. T. Luning Prak, P. Bates, S. E. Hensley, A. R. Greenplate, E. J. Wherry, Distinct antibody and memory B cell responses in SARS-CoV-2 naïve and recovered individuals following mRNA vaccination. Sci Immunol 6, (2021).

23. Y. Liu, H. X. Tan, M. Koutsakos, S. Jegaskanda, R. Esterbauer, D. Tilmanis, M. Aban, K. Kedzierska, A. C. Hurt, S. J. Kent, A. K. Wheatley, Cross-lineage protection by human antibodies binding the influenza B hemagglutinin. Nat Commun 10, 324 (2019).

24. S. H. Wilks, B. Muhlemann, X. Shen, S. Tureli, E. B. LeGresley, A. Netzl, M. A. Caniza, J. N. Chacaltana-Huarcaya, V. M. Corman, X. Daniell, M. B. Datto, F. S. Dawood, T. N. Denny, C. Drosten, R. A. M. Fouchier, P. J. Garcia, P. J. Halfmann, A. Jassem, L. M. Jeworowski, T. C. Jones, Y. Kawaoka, F. Krammer, C. McDanal, R. Pajon, V. Simon, M. S. Stockwell, H. Tang, H. van Bakel, V. Veguilla, R. Webby, D. C. Montefiori, D. J. Smith, Mapping SARS-CoV-2 antigenic relationships and serological responses. Science 382, eadj0070 (2023).

25. S. H. Wilks, titertools. https://github.com/shwilks/titertools, (2023).

26. W. Zhang, L. C. Rowntree, R. Muttucumaru, T. Damelang, M. Aban, A. C. Hurt, M. Auladell, R. Esterbauer, B. Wines, M. Hogarth, S. J. Turner, A. K. Wheatley, S. J. Kent, S. Patil, S. Avery, O. Morrissey, A. W. Chung, M. Koutsakos, T. H. Nguyen, A. C. Cheng, T. C. Kotsimbos, K. Kedzierska, Robust immunity to influenza vaccination in haematopoietic stem cell transplant recipients following reconstitution of humoral and adaptive immunity. Clin Transl Immunology 12, e1456 (2023).

27. Selva, K. Selva, C. E. van de Sandt, M. Lemke, C. Lee, S. Shoffner, B. Chua, S. Davis, T. H. O. Nguyen, L. Rowntree, L. Hensen, M. Koutsakos, C. Wong, F. Mordant, D. Jackson, K. Flanagan, J. Crowe, S. Tosif, M. Neeland, P. Sutton, P. Licciardi, N. Crawford, A. Cheng, D. Doolan, F. Amanat, F. Krammer, K. Chappell, N. Modhiran, D. Watterson, P. Young, W. Lee, B. Wines, P. Mark Hogarth, R. Esterbauer, H. Kelly, H.-X. Tan, J. Juno, A. Wheatley, S. Kent, K. Arnold, K. Kedzierska, A. W. Chung, Systems serology detects functionally distinct coronavirus antibody features in children and elderly. Nature Communications 12, 2037 (2021).

28. J. R. Habel, B. Y. Chua, L. Kedzierski, K. J. Selva, T. Damelang, E. R. Haycroft, T. H. Nguyen, H. F. Koay, S. Nicholson, H. A. McQuilten, X. Jia, L. F. Allen, L. Hensen, W. Zhang, C. E. van de Sandt, J. A. Neil, K. Pragastis, J. S. Lau, J. Jumarang, E. K. Allen, F. Amanant, F. Krammer, K. M. Wragg, J. A. Juno, A. K. Wheatley, H. X. Tan, G. Pell, S. Walker, J. Audsley, A. Reynaldi, I. Thevarajan, J. T. Denholm, K. Subbarao, M. P. Davenport, P. M. Hogarth, D. I. Godfrey, A. C. Cheng, S. Y. Tong, K. Bond, D. A. Williamson, J. H. McMahon, P. G. Thomas, P. S. Pannaraj, F. James, N. E. Holmes, O. C. Smibert, J. A. Trubiano, C. L. Gordon, A. W. Chung, C. L. Whitehead, S. J. Kent, M. Lappas, L. C. Rowntree, K. Kedzierska, Immune profiling of SARS-CoV-2 infection during pregnancy reveals NK cell and γδ T cell perturbations. JCI Insight 8, (2023).

