## Supplementary data for "Historic 1994 influenza vaccine cohorts reveal breadth of antibody and B cell responses towards three decades of future influenza A and B viruses"

**List of Figures**

**Figure S1.** Validation of multiplex bead array assay. Related to Fig. 4.
**Figure S2.** H1N1 responses across IgG subtypes. Related to Fig. 4.
**Figure S3.** H1N1 responses across Fcγ receptors. Related to Fig. 4.
**Figure S4.** H3N2 responses across IgG subtypes and Fcγ receptors. Related to Fig. 4.
**Figure S5.** FLUBV responses IgG subtypes and Fcγ receptors. Related to Fig. 4.
**Figure S6.** Volcano plots of antigen-antibody features from systems serology analyses. Related to Fig. 4.
**Figure S7.** PCA analyses of young and older adults.
**Figure S8.** FACS gating strategies to measure probe-specific memory B cell responses. Related to Fig. 5 and 6.
**Figure S9.** Validation of recombinant H1, H3, and IBV HA probes on donors who received the IIV in 2022. Related to Fig. 5.
**Figure S10.** Phenotype and isotype of future virus strain-specific memory B cell responses. Related to Fig. 5 and 6.

**List of Tables**

**Table S1.** List of influenza virus strains used in the study. Related to Fig. 2 and 3.
**Table S2.** Influenza antigens used for the multiplex assay. Related to Fig. 4.
**Table S3.** Fluorescently labelled recombinant HA-specific probes. Related to Fig. 5 and 6.
**Table S4.** Mouse anti-human antibody detectors used in multiplex assay. Related to Fig. 4.
**Table S5.** Mouse anti-human antibody panel for measuring HA-specific B cells. Related to Fig. 5 and 6.

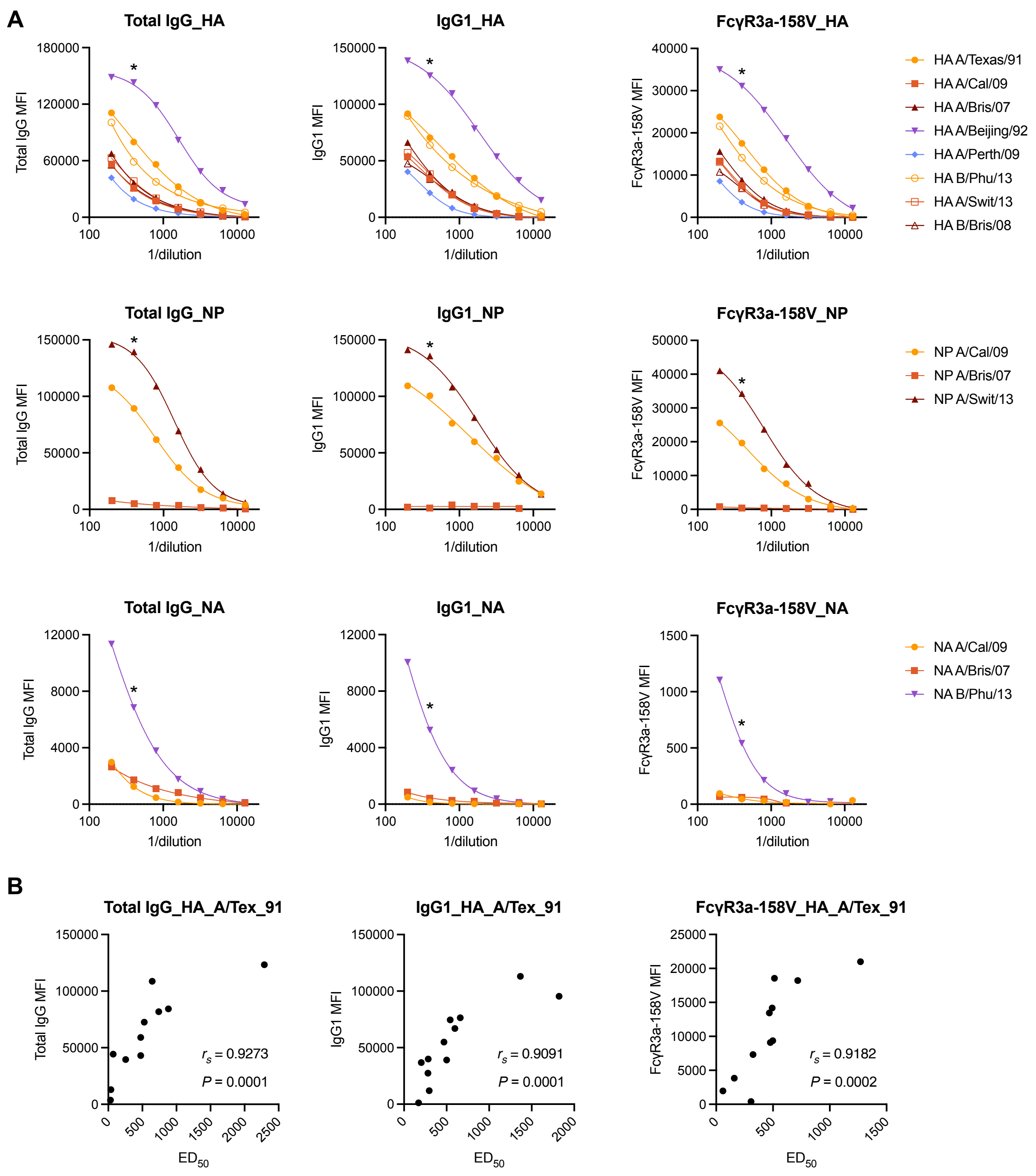

**Figure S1.** Validation of multiplex bead array assay. (A) Representative dilution curves depicting total IgG, IgG1, and FcgR3a-131V responses of pooled plasma against the HA, NP, and NA influenza antigens included in the multiplex panel. Single asterisks indicate the selected 1:400 dilution. (B) Representative Spearman correlations of MFIs for the selected 1:400 dilution versus $ED_{50}$s against the vaccine antigen HA A/Tex/91 for the individual samples included in the plasma pool. Strong correlation of the single dilution against the $ED_{50}$ indicates that the selected dilution is appropriate for assessment of MFIs across the range of tested responses. Related to Fig. 4.

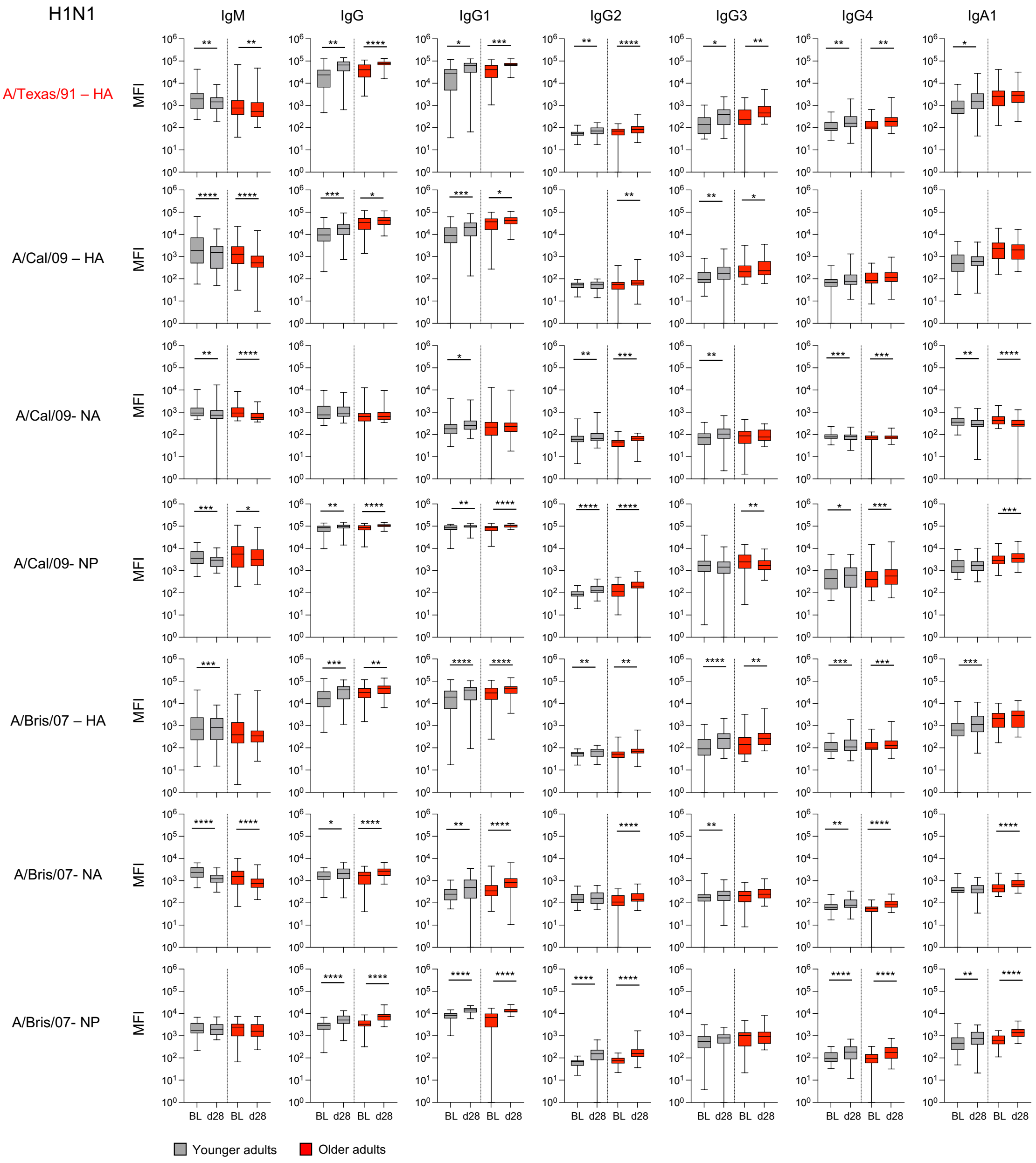

**Figure S2.** H1N1 responses across IgG subtypes. MFIs are shown for each antigen per detector antibody. The bounds of the box plot indicate the 25th and 75th percentiles, the median as the central bar, and the whiskers represent the minimum and maximum values. Statistical significance determined by Wilcoxon test for timepoint comparisons. *$P < 0.05$, **$P < 0.01$, ***$P < 0.001$, ****$P < 0.0001$. Related to Fig. 4.

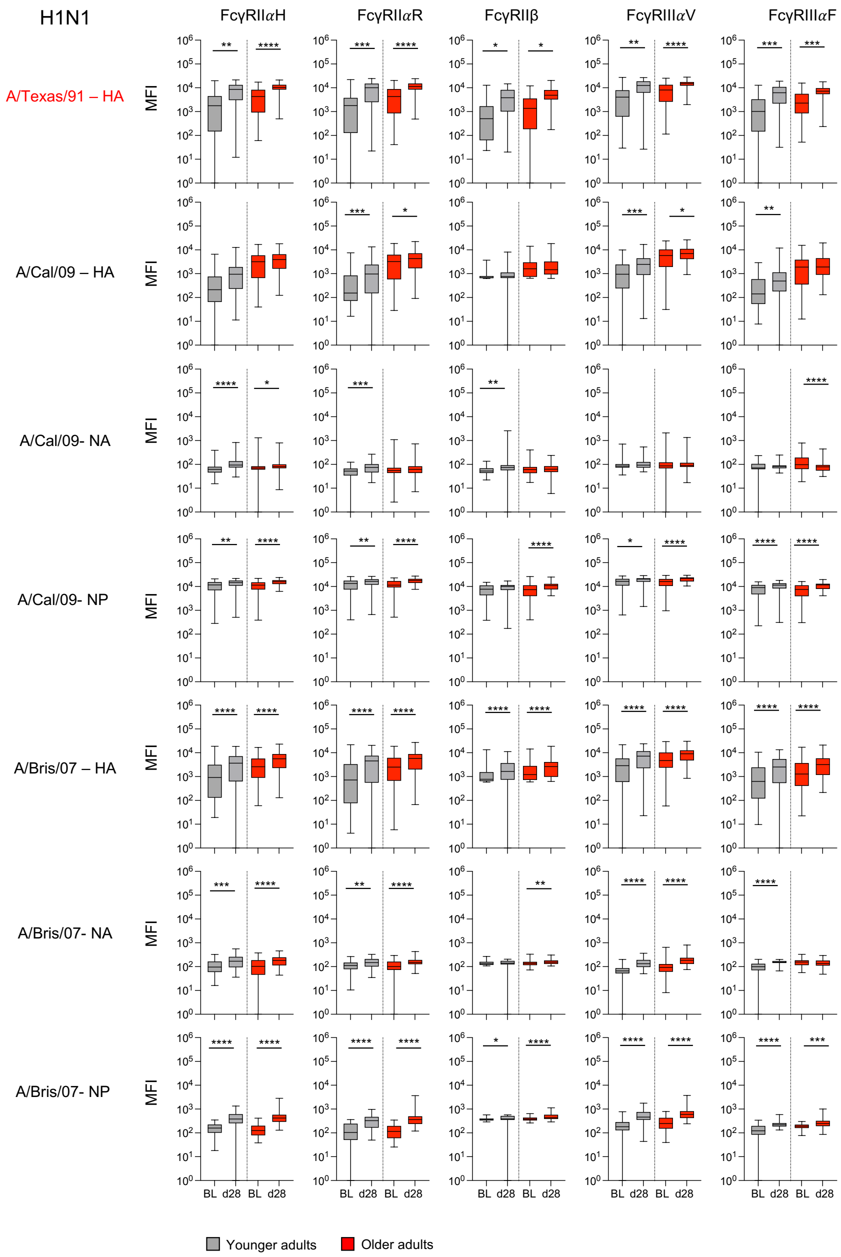

**Figure S3.** H1N1 responses across Fcγ receptors. MFIs are shown for each antigen per detector antibody. The bounds of the box plot indicate the 25th and 75th percentiles, the median as the central bar, and the whiskers represent the minimum and maximum values. Statistical significance determined by Wilcoxon test for timepoint comparisons. *$P < 0.05$, **$P < 0.01$, ***$P < 0.001$, ****$P < 0.0001$. Related to Fig. 4.

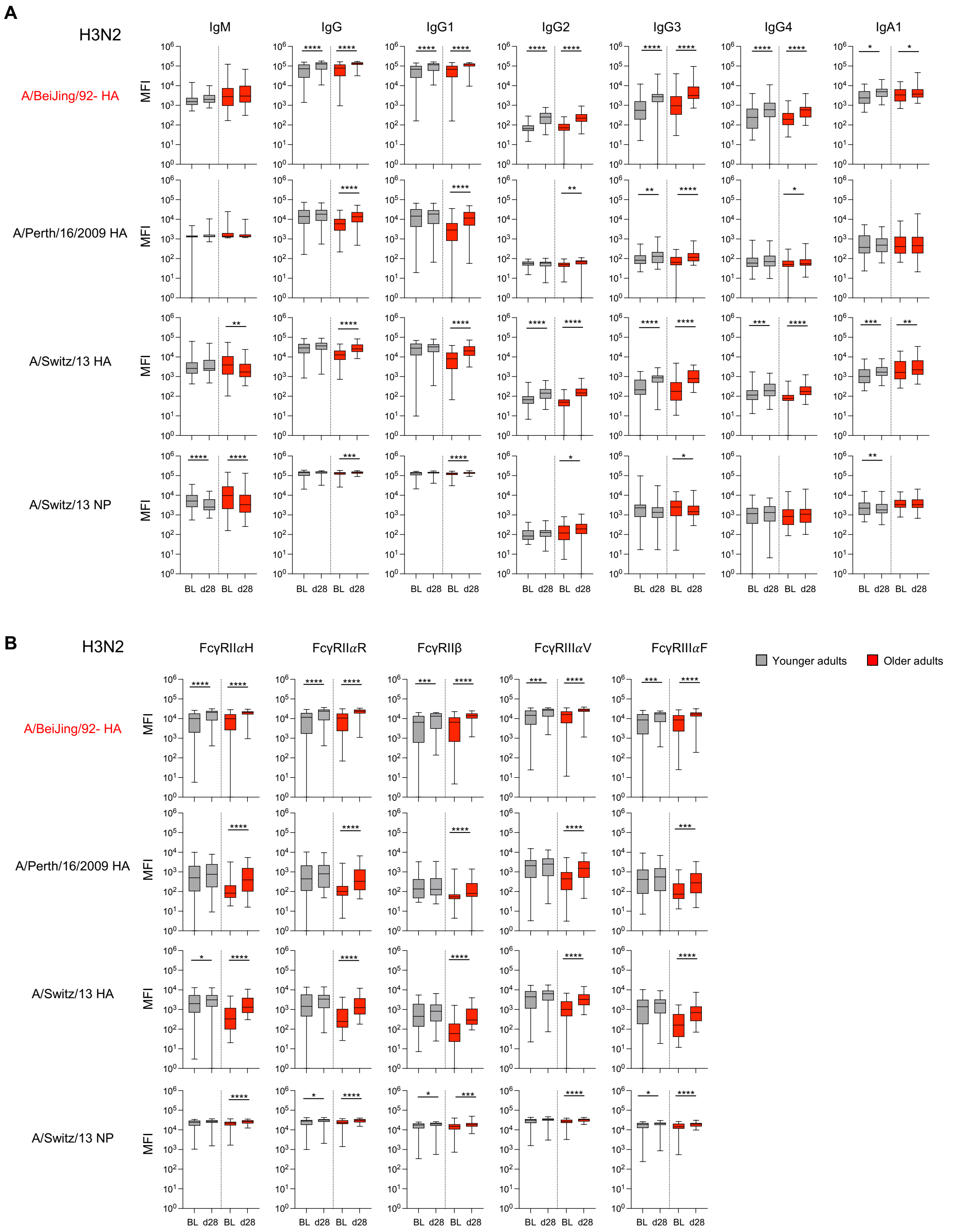

**Figure S4.** H3N2 responses across IgG subtypes and Fcγ receptors. MFIs are shown for each antigen per detector antibody. The bounds of the box plot indicate the 25th and 75th percentiles, the median as the central bar, and the whiskers represent the minimum and maximum values. Statistical significance determined by Wilcoxon test for timepoint comparisons. **P* < 0.05, ***P* < 0.01, ****P* < 0.001, *****P* < 0.0001. Related to Fig. 4.

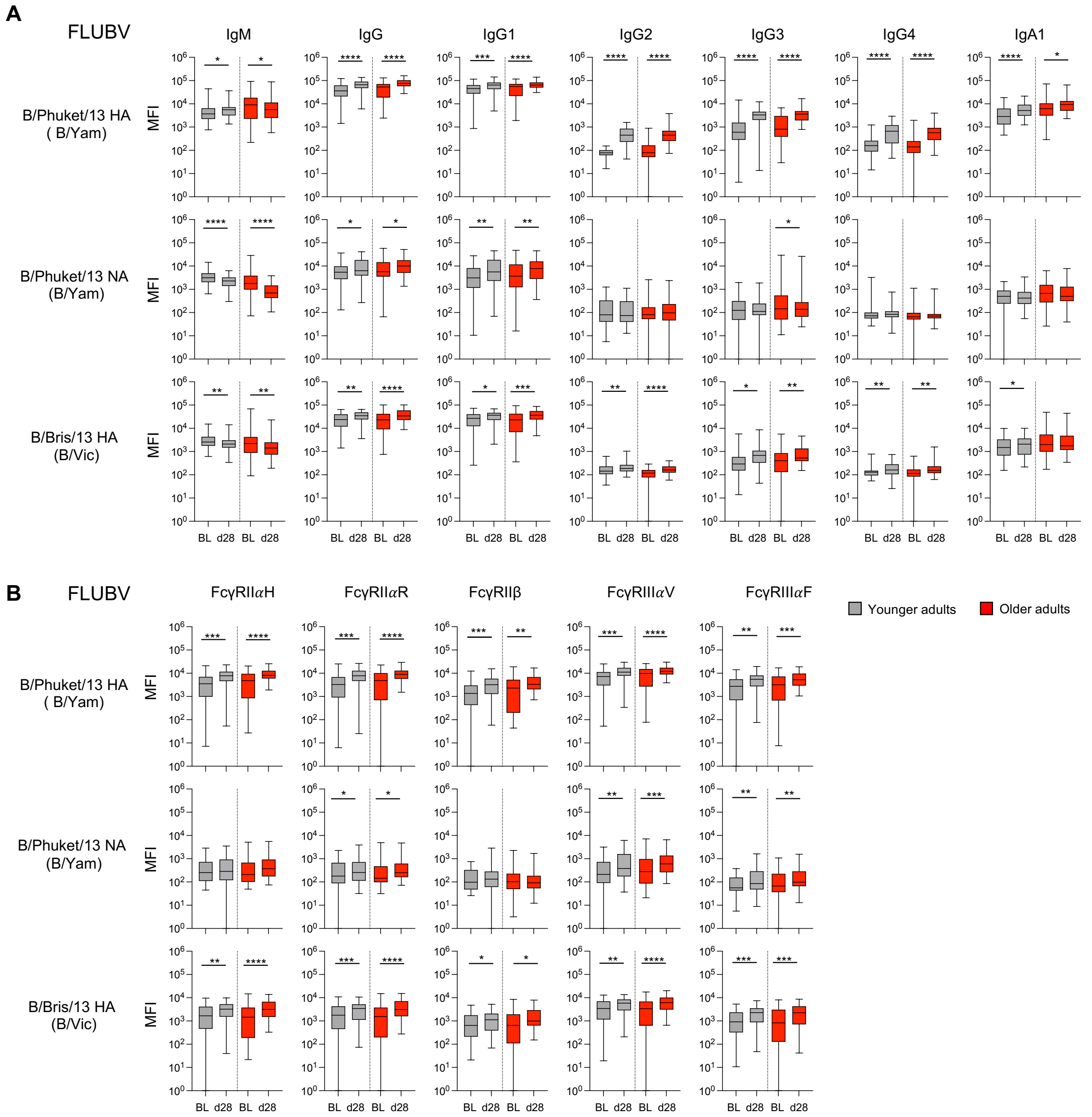

**Figure S5.** FLUBV responses IgG subtypes and Fcγ receptors. MFIs are shown for each antigen per detector antibody. The bounds of the box plot indicate the 25th and 75th percentiles, the median as the central bar, and the whiskers represent the minimum and maximum values. Statistical significance determined by Wilcoxon test for timepoint comparisons. *$P < 0.05$, **$P < 0.01$, ***$P < 0.001$, ****$P < 0.0001$. Related to Fig. 4.

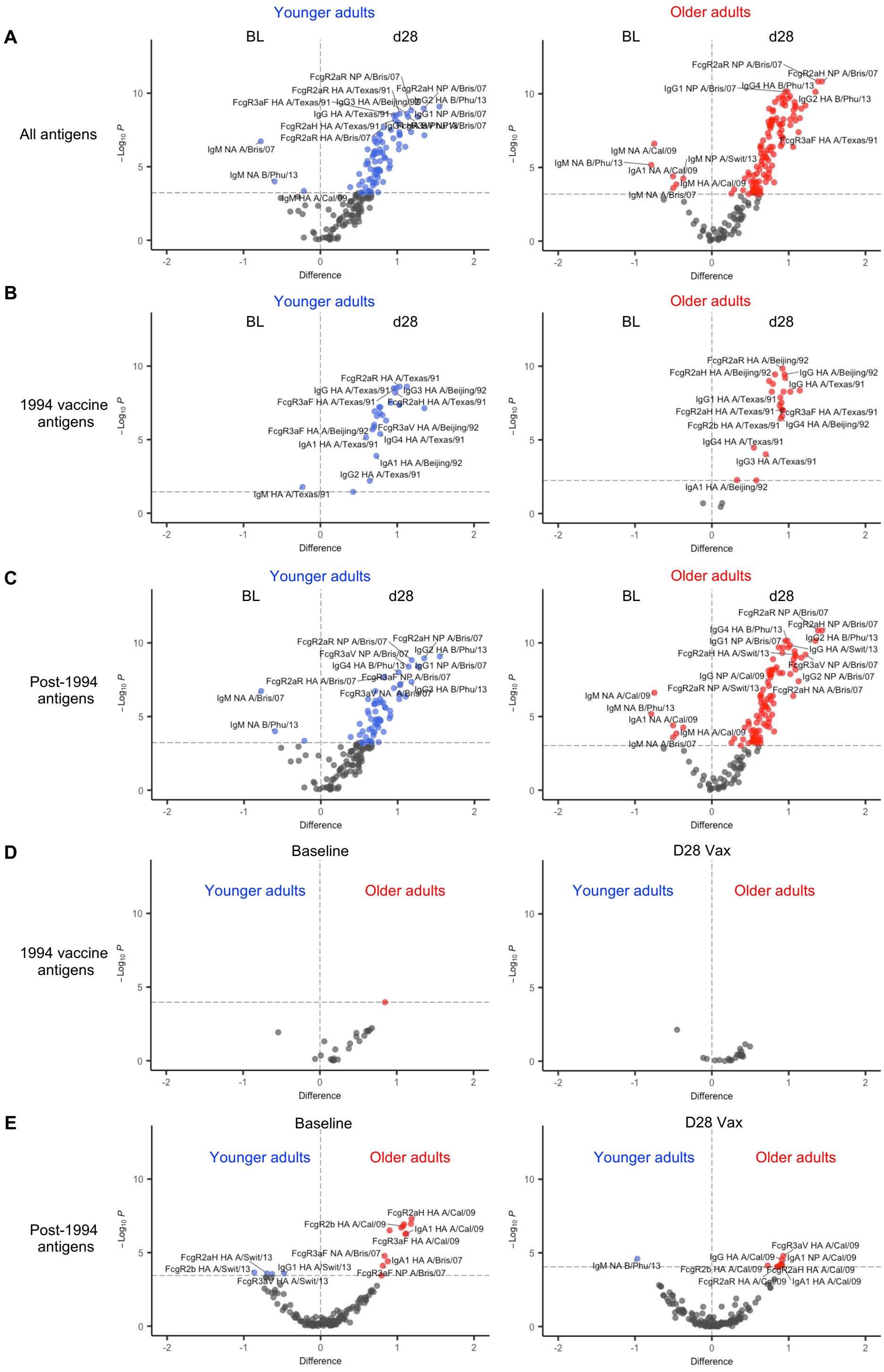

**Figure S6.** Volcano plots of antigen-antibody features from systems serology. Volcano plots between baseline and d28 post vaccination for young and older adults for (A) all antigens, (B) 1994 vaccine antigens or (C) post 1994 antigens. Volcano plots between young and older adults at baseline and d28 post vaccination for (D) 1994 vaccine antigens or (E) post 1994 antigens. 1994 vaccine antigens included H1-HA A/Texas/36/1991 and H3-HA A/Beijing/32/1992. Z-score differences in $Log_{10}$(MFI+1) for each variable from baseline to d28 post vaccination were determined by Wilcoxon matched-pairs signed rank test, and between age cohorts by Wilcoxon rank-sum/Mann-Whitney U test. The Holm correction for multiple comparisons was made; unadjusted $-Log_{10}$ $P$ values are plotted, with the horizontal threshold line plotted at an adjusted ($-Log_{10}$) $P$ value of 0.05. Related to Fig. 4.

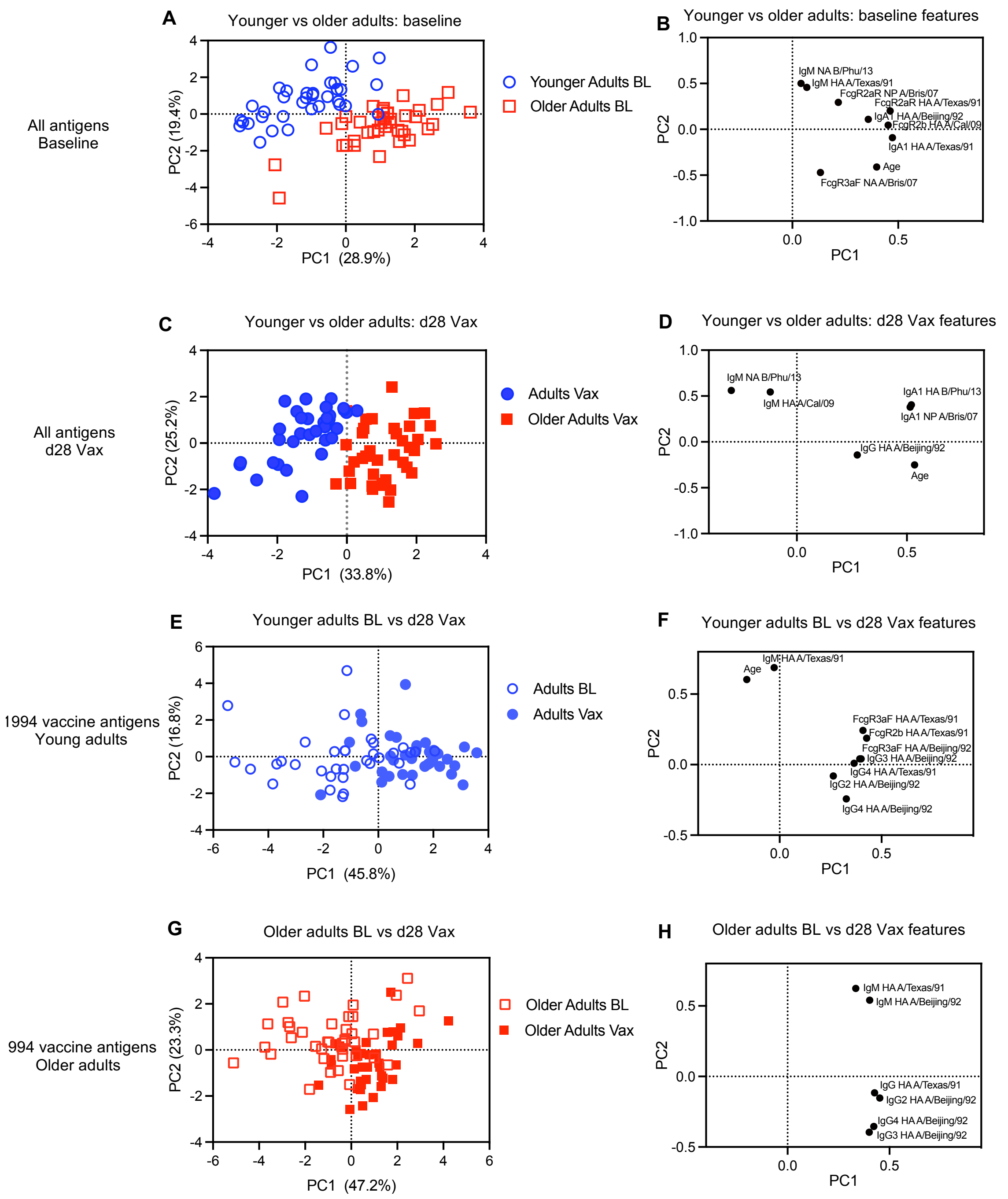

**Figure S7.** PCA analyses of young and older adults. PCA of feature selected data from all antigens including age as the variable features comparing (A) antibody responses for young and older adults at baseline with (B) PC1 of antibody features and at (C) d28 post vaccination with (D) PC1 of selected features. (E) PCA scores and (F) PC1 of antibody features from 1994 vaccine antigens plus age as variable features comparing young adults at pre (BL) and post vaccination (d28 Vax), and (G,H) comparing older adults at pre and post vaccination. 1994 vaccine antigens included H1-HA A/Texas/36/1991 and H3-HA A/Beijing/32/1992. Related to Fig 4.

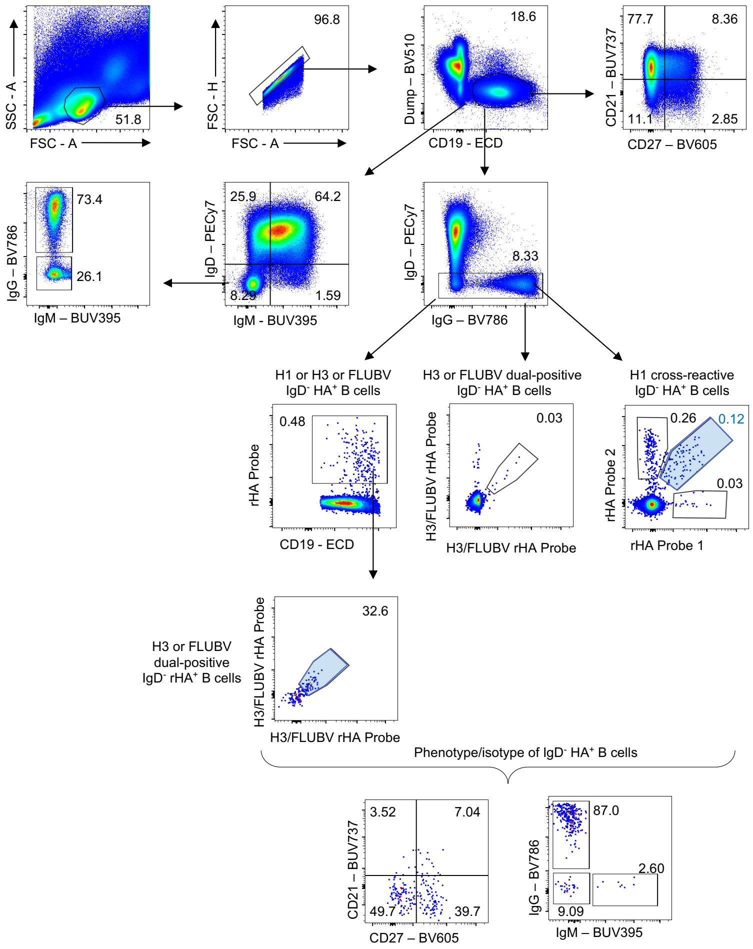

**Figure S8.** Gating strategy for HA-specific memory B cells. Memory B cells were gated as singlets, then dump$^-$CD19$^+$IgD$^-$ cells before single and dual HA-probe gates. Phenotype gates (CD21 versus CD27) and isotype gates (IgG versus IgM) are shown for total CD19$^+$ B cells and IgD$^-$HA-probe$^+$ B cells. Related to Fig. 5 and 6.

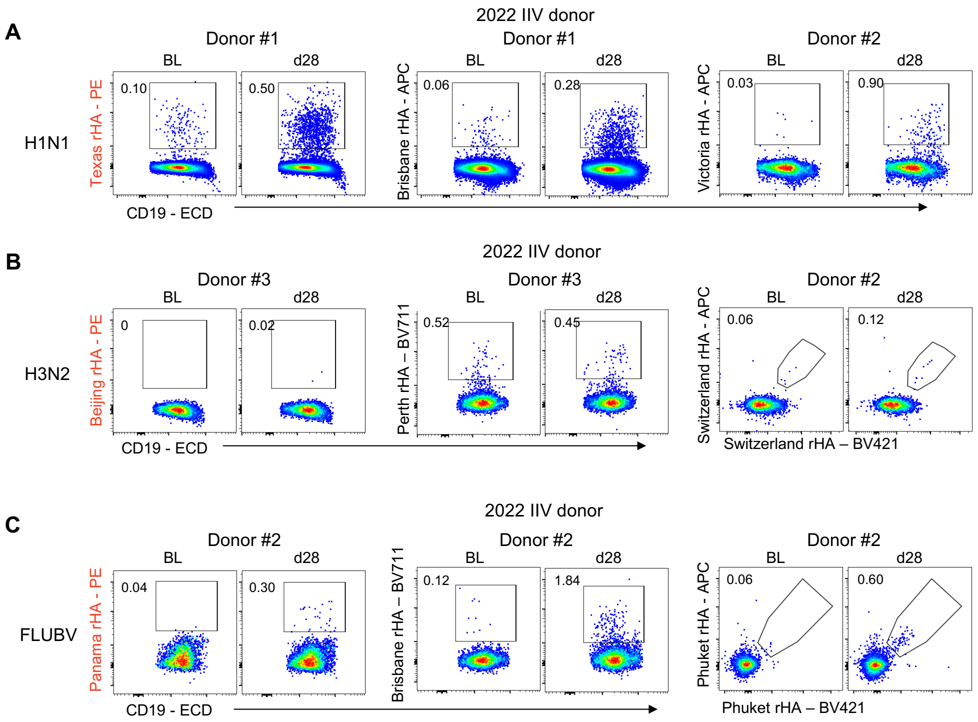

**Figure S9.** Validation of recombinant H1, H3, and IBV HA probes. Representative HA-probe staining for 1994 IIV strain-specific memory B cells on donors vaccinated in 2022 with the IIV. Representative staining of (A) H1- (B) H3- and (C) FLUBV-specific B cell responses (n = 5-15).

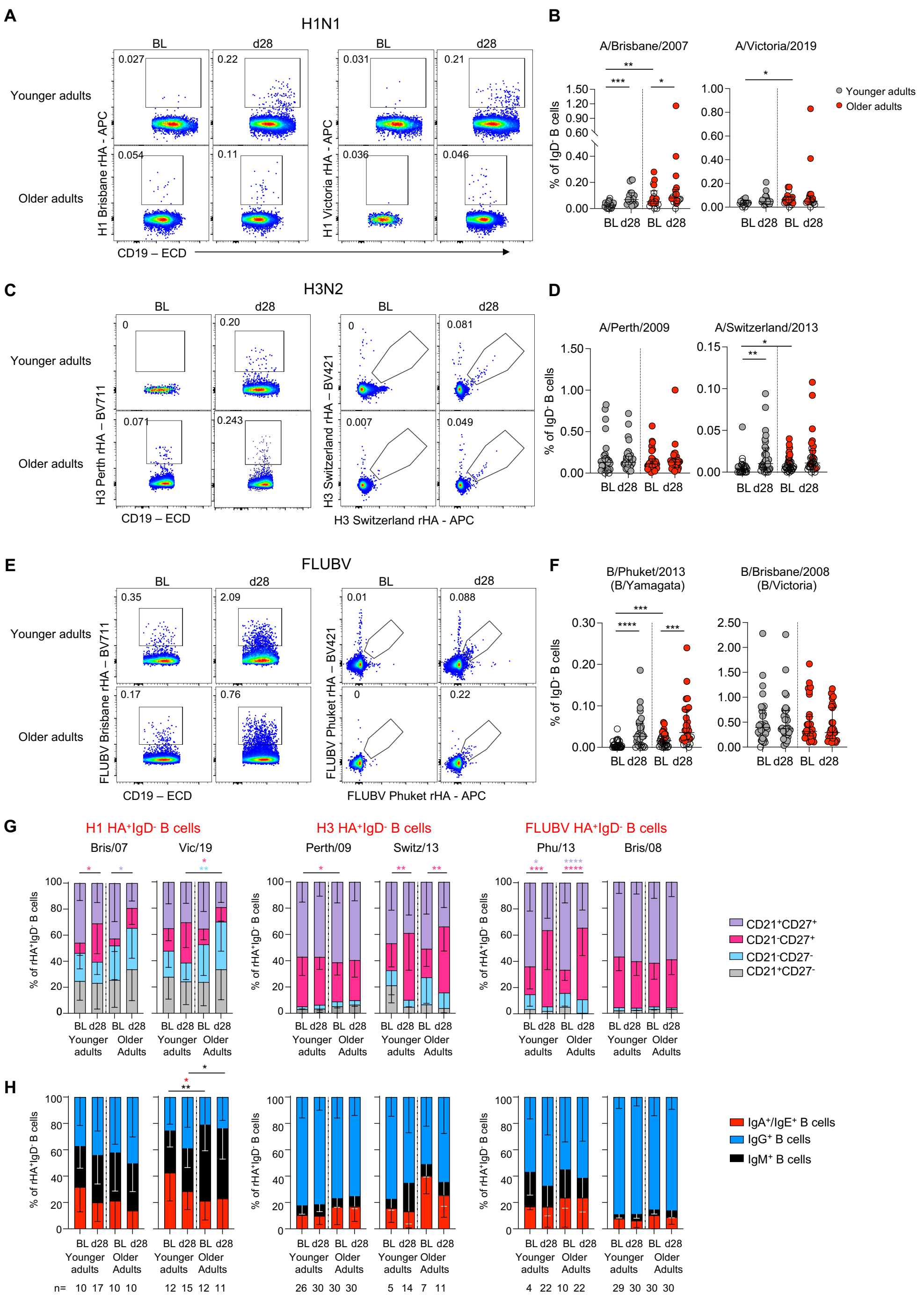

**Figure S10.** Phenotype and isotype of future virus strain-specific memory B cell responses. (A) Representative HA-probe staining for H1 future strain-specific memory B cells. (B) Frequency of future H1 strain-specific IgD- B cells at baseline and d28 post vaccination in adults and older adults (Young adults n=19, Older adults n=19). (C) Representative staining and (D) frequency of future H3 strain-specific B cell responses (Young adults n=30, Older adults n=30). (E) Representative staining and (F) frequency of future FLUBV-specific B cell responses (Young adults n=30, Older adults n=30). Medians and IQRs are shown. Statistical significance determined by Wilcoxon test for timepoint comparisons within an age group or by Mann-Whitney for comparisons between adults and older adults. (G) Phenotype and (H) isotype of future strain-specific memory B cells. Mean and SD are shown. Statistical significance determined by Tukey's multiple comparisons test. *$P < 0.05$, **$P < 0.01$, ***$P < 0.001$, ****$P < 0.0001$. Related to Fig. 5 and 6.

**Table S1.** List of influenza virus strains used in the study. Related to Fig. 2 and 3.

| Subtype | Strain | Source | Years included in Australian IIV since 1990 |
|---|---|---|---|
| A/H1N1 | A/Puerto Rico/08/1934 | CSL Seqirus Ltd | - |
| A/H1N1 | A/Brazil/11/1978 | WHO FLU[2] | - |
| A/H1N1 | A/Texas/36/1991[1] | WHO FLU | 1993-1997 |
| A/H1N1 | A/Beijing/262/1995 | WHO FLU | 1999 |
| A/H1N1 | A/New Caledonia/20/1999 | WHO FLU | 2000-2007 |
| A/H1N1 | A/Solomon Islands/3/2006 | CSL Seqirus Ltd | 2008 |
| A/H1N1 | A/Brisbane/59/2007 | CSL Seqirus Ltd | 2009 |
| A/H1N1 | A/California/7/2009 | CSL Seqirus Ltd | 2010-2016 |
| A/H1N1 | A/Michigan/45/2015 | CSL Seqirus Ltd | 2017-2019 |
| A/H3N2 | A/Port Chalmers/1/1973 | WHO FLU | - |
| A/H3N2 | A/Beijing/32/1992[1] | WHO FLU | 1994 |
| A/H3N2 | A/Wuhan/359/1995 | WHO FLU | 1997 |
| A/H3N2 | A/Sydney/5/1997 | WHO FLU | 1998-2000 |
| A/H3N2 | A/California/7/2004 | WHO FLU | 2006 |
| A/H3N2 | A/Wisconsin/67/2005 | WHO FLU & CSL Seqirus Ltd | 2007 |
| A/H3N2 | A/Brisbane/10/2007 | CSL Seqirus Ltd | 2008-2009 |
| A/H3N2 | A/Perth/16/2009 | CSL Seqirus Ltd | 2010-2012 |
| A/H3N2 | A/Victoria/361/2011 | CSL Seqirus Ltd | 2013 |
| A/H3N2 | A/Texas/50/2012 | CSL Seqirus Ltd | 2014 |
| A/H3N2 | A/Switzerland/9715292/2013 | CSL Seqirus Ltd | 2015 |
| A/H3N2 | A/Hong Kong/4801/2014 | CSL Seqirus Ltd | 2016-2017 |
| A/H3N2 | A/Sinagpore/16/2016 | CSL Seqirus Ltd | 2018 |
| A/H3N2 | A/Switzerland/8060/2017 | CSL Seqirus Ltd | 2019 |
| B/Yamagata | B/Yamagata/16/1988 | WHO FLU | 1990-1992 |
| B/Yamagata | B/Panama/45/1990[1] | WHO FLU | 1992-1995 |
| B/Yamagata | B/Florida/4/2006 | CSL Seqirus Ltd | 2008-2009 |
| B/Yamagata | B/Wisconsin/1/2010 | CSL Seqirus Ltd | 2013 |
| B/Yamagata | B/Massachusetts/2/2012 | CSL Seqirus Ltd | 2014 |
| B/Yamagata | B/Phuket/3073/2013 | CSL Seqirus Ltd | 2015, 2018-2025 |
| B/Victoria | B/Victoria/02/1987 | WHO FLU | - |
| B/Victoria | B/Malaysia/2506/2004 | CSL Seqirus Ltd | 2006-2007 |
| B/Victoria | B/Brisbane/60/2008 | CSL Seqirus Ltd | 2010-2012, 2016-2017 |

[1]Influenza virus strains in red were included in the 1994 Fluvax.

[2]WHO FLU = WHO Collaborating Centre for Reference and Research on Influenza (Melbourne, Australia).

**Table S2.** Influenza antigens used for the multiplex assay. 1994 Vaccine antigens shown in red. Related to Fig. 4.

| Antigen | Subtype | Strain | µg /1.25e7 beads | Source | Catalogue number |
|---|---|---|---|---|---|
| HA | H1N1 | A/Texas/36/1991 | 54 | Sino | 40713-V08H |
| HA | H1N1 | A/Brisbane/59/2007 | 59 | Sino | 11052-V08H |
| HA | H1N1 | A/California/07/2009 | 55 | Sino | 11055-V08H |
| HA | H3N2 | A/Beijing/32/1992 | 70 | Immune Tech | IT-003-00411ΔTMp |
| HA | H3N2 | A/Perth/16/2009 | 55 | Sino | 40043-V08H |
| HA | H3N2 | A/Switzerland/9715293/2013 | 60 | Sino | 40497-V08B |
| HA | B/Vic | B/Brisbane/60/2008 | 56 | Bei Resource | NR-56334 |
| HA | B/Yam | B/Phuket/3073/2013 | 55 | Sino | 40498-V08B |
| NA | H1N1 | A/Brisbane/59/2007 | 48 | Bei Resource | NR-43785 |
| NA | H1N1 | A/California/07/2009 | 53 | Sino | 11058-VNAHC1 |
| NA | B/Yam | B/Phuket/3073/2013 | 53 | Sino | 40502-V07B |
| NP | H1N1 | A/Brisbane/59/2007 | 27 | Immune Tech | IT-003-028Ep |
| NP | H1N1 | A/California/07/2009 | 27 | Sino | 40205-V08B |
| NP | H3N2 | A/Switzerland/9715293/2013 | 28 | Sino | 40499-V08B |
| SIVgp120 | N/A | Negative antigen control | 100 | Sino | 40415-V08H |
| Tetanus toxoid | N/A | Positive antigen control | 56 | Sigma-Aldrich | T3194-25UG |

**Table S3.** Fluorescently labelled recombinant HA-specific probes. Related to Fig. 5 and 6.

| **Panel** | **Subtype** | **Strain** | **Fluorochrome** | **Source** | **PMID** |
|---|---|---|---|---|---|
| 1 | H1N1 | A/Texas/36/1991 | PE | Produced in-house | |
| 1 | H1N1 | A/Brisbane/59/2007 | APC | Produced in-house | |
| 1 | H1N1 | A/Victoria/2570/2019 (H1N1)pdm09-like | APC | Produced in-house | 40600710 |
| 2 | H3N2 | A/Beijing/32/1992 | PE | Produced in-house | |
| 2 | H3N2 | A/Perth/16/2009 | BV711 | Produced in-house | |
| 2 | H3N2 | A/Switzerland/9715292/2013 | APC, BV421 | Produced in-house | 33976217 |
| 3 | B/Yam | B/Panama/45/1990 | PE | Produced in-house | |
| 3 | B/Vic | B/Brisbane/60/2008 | BV711 | Produced in-house | 33976217 |
| 3 | B/Yam | B/Phuket/3073/2013 | APC, BV421 | Produced in-house | 33976217 |

**Table S4.** Mouse anti-human antibody detectors used in multiplex assay. Related to Fig. 4.

| **Detector, PE** | **Clone** | **Source** | **Catalogue number** |
|---|---|---|---|
| IgA1 | B3506B4 | Southern Biotech | 9130-09 |
| IgM | SA-DA4 | Southern Biotech | 9020-09 |
| Total IgG | JDC-10 | Southern Biotech | 9040-09 |
| IgG1 | HP6001 | Southern Biotech | 9054-09 |
| IgG4 | HP6025 | Southern Biotech | 9200-09 |
| **Detector, biotin** | **Clone** | **Source** | **Catalogue number** |
| IgG2 | HP6200 | MabTech | 3852-6-250 |
| IgG3 | MTG34 | MabTech | 3853-6-250 |
| FcγRIIa-H131 | N/A | Produced in-house | PMID: 27385782 |
| FcγRIIa-R131 | N/A | Produced in-house | PMID: 27385782 |
| FcγRIIb | N/A | Produced in-house | PMID: 40527060 |
| FcγRIIIa-V158 | N/A | Produced in-house | PMID: 27385782 |
| FcγRIIIa-F158 | N/A | Produced in-house | PMID: 27385782 |
| Streptavidin R-Phycoerythrin Conjugate | N/A | Thermo Fisher | S866 |

**Table S5.** Mouse anti-human antibody panel for measuring HA-specific B cells. Related to Fig. 5 and 6.

| **Antibody** | **Clone** | **Dilution** | **Company** | **Catalog** |
|---|---|---|---|---|
| IgM BUV395 | G20-127 | 1:150 | BD | 563903 |
| CD21 BUV737 | B-ly4 | 1:300 | BD | 612788 |
| Streptavidin BV510 | N/A | 1:600 | BD | 563261 |
| LIVE/DEAD Aqua | N/A | 1:500 | Thermofisher | L34966 |
| CD3 BV510 | OKT3 | 1:600 | Biolegend | 317332 |
| CD8 BV510 | RPA-T8 | 1:1500 | Biolegend | 301047 |
| CD10 BV510 | HI10a | 1:750 | Biolegend | 312219 |
| CD14 BV510 | M5E2 | 1:300 | Biolegend | 301841 |
| CD16 BV510 | 3G8 | 1:500 | Biolegend | 302047 |
| CD27 BV605 | O323 | 1:150 | Biolegend | 302830 |
| CD71 BV650 | CY1G4 | 1:75 | Biolegend | 334116 |
| IgG BV786 | G18-145 | 1:75 | BD | 564230 |
| CD20 AF700 | 2H7 | 1:150 | BD | 560631 |
| CD11c FITC | Bu15 | 1:50 | Biolegend | 337214 |
| CD19 ECD | J3-119 | 1:150 | Beckman | IM2708U |
| IgD PE-Cy7 | IA6-2 | 1:500 | BD | 561314 |
